# Gastric digestion and changes in serum amino acid concentrations after consumption of casein from cow and goat milk: a randomized crossover trial in healthy men

**DOI:** 10.1101/2024.04.10.24305606

**Authors:** Elise J.M. van Eijnatten, Guido Camps, Wolf Rombouts, Linette Pellis, Paul A.M. Smeets

## Abstract

**Background:** *In vitro* studies show that goat milk proteins form less compact coagulates in the stomach compared to cow milk proteins, which may facilitate gastric digestion and amino acid (AA) absorption. However, this has not been confirmed *in vivo* in humans.

**Objective:** To examine gastric digestion and changes in AA concentrations after cow milk-derived (cow MC) and goat milk-derived casein (goat MC) ingestion.

**Methods:** In this single-blind randomized cross-over study 18 men (age 23 ± 1.6 years, BMI 23 ± 1.6 kg/m^2^) consumed 300 ml of a drink containing 30 g of cow MC or goat MC. Participants underwent gastric MRI scans at baseline and every 10 minutes up to 60 minutes postprandially. Blood was drawn at baseline and up to 4 hours postprandially. In addition, participants verbally rated their appetite after each MRI measurement. Primary outcomes were gastric emptying and AA concentrations. Secondary outcome was gastric coagulation as inferred by image texture metrics.

**Results:** Gastric emptying half-time was 80 ± 25 minutes for goat and 85 ± 24 minutes for cow MC (p = 0.395). In line with this, gastric emptying of the drinks over time was similar (MD 0.77 ml, 95% CI [-6.9, 8.5], p=0.845). Serum essential AA (MD -110 µmol/L, 95% CI [-162, -58]) was higher over time for cow MC (p<0.001). The image texture metric contrast was lower for cow MC (MD 0.010, 95% CI [0.001, 0.020], p=0.036).

**Conclusion:** Cow MC and goat MC have different coagulating properties, as inferred by AA concentrations and supported by image texture analysis. This did not influence overall gastric emptying or the emptying of the liquid and coagulated fractions, which were similar. This warrants further *in vivo* research on casein coagulation in the food matrix to help determine the optimal use for cow and goat milk and their protein fractions.

Financial support: the study was funded by Ausnutria Dairy Corporation Ltd.

Clinical trial registry number: NL8137 (Netherlands Trial Registry), accessible through https://trialsearch.who.int/Trial2.aspx?TrialID=NL-OMON28580

## INTRODUCTION

Protein is an essential macronutrient used in many processes in the human body (Atherton and Smith 2012, Do, Lewis et al. 2019). It is important that ingested protein is properly digested and absorbed so that it can be used for protein synthesis (Mahé, Roos et al. 1996, van Vliet, Burd et al. 2015, Fardet, Dupont et al. 2019). Dairy products constitute a significant protein source globally (Lagrange, Whitsett et al. 2015). Cow milk dairy products are the most commonly used, but goat milk popularity is increasing (Miller and Lu 2019). One of the reasons for this is the consumers’ perception of its health benefits. These benefits are hypothesized to originate from a difference in the milks’ digestion due to their different casein composition (Roy, Moughan et al. 2022).

Cow and goat milk generally contain about 3.5 % protein of which caseins represent about 80% and whey proteins about 20% (Ceballos, Morales et al. 2009). During digestion in the stomach the casein micelles (CM) are destabilized by pepsin proteolysis and acidification. This results in the formation of coagulates containing protein, and if present, fat globules (Ye, Liu et al. 2019). The physical properties of these casein coagulates could affect gastric protein digestion, gastric emptying and subsequent intestinal digestion and absorption of amino acids (AA) (Huppertz and Chia 2021). Previous studies, predominantly *in vitro*, have shown that casein coagulation is affected by several factors including processing-induced protein modifications, overall product composition (food matrix), and differences in protein composition for instance between animal species (Almaas, Cases et al. 2006, Ceballos, Morales et al. 2009, Wang, Ye et al. 2018, Mulet-Cabero, Mackie et al. 2019, Eijnatten, Roelofs et al. 2023, Hettinga, Pellis et al. 2023). Human *in vivo* studies on protein modifications, resulting in different cow casein forms, showed that coagulation and gastric emptying can have strong effects on the postprandial rise in AA bioavailability (Trommelen, Weijzen et al. 2020). Even though goat micellar casein seemed to have similar protein structures on a molecular level in an *in vitro* study using X-ray scattering (Ingham, Smialowska et al. 2018), casein micelles differ in size, hydration, and mineralization compared to bovine caseins (Claeys, Verraes et al. 2014, Roy, Moughan et al. 2022). During static *in vitro* digestion, more β-casein and less αs1-casein may contribute to the formation of looser gastric clots in goat casein micelles and therefore greater proteolysis than cow casein micelles (Zhang, Liu et al. 2023). Increased access of the enzymes to the proteins might lead to faster gastric digestion of goat as compared to cow milk-derived caseins and milk, as seen in *in vitro* coagulum analysis (Park 2017). In line with this, infant formula based on goat milk formed smaller flocs of aggregated protein and oil droplets under gastric conditions, leading to faster protein digestion in goat milk infant formula than seen in cow milk based infant formula (Ye, Cui et al. 2019). Similarly, goat milk proteins were digested faster than cow milk proteins in *in vitro* digestion by human gastric and duodenal enzymes (Almaas, Cases et al. 2006). This was also seen in two *in vitro* studies under simulated infant conditions where proteins in goat milk and goat milk-based infant formula had different digestive behaviour compared to those present in cow milk and cow milk-based formula (Maathuis, Havenaar et al. 2017, Hodgkinson, Wallace et al. 2018). Maathuis et al. controlled for gastric emptying. Therefore differences in digestion were probably due to differences in coagulation. However, not all studies showed a faster digestion of goat milk protein. Inglingstad et al. found in an *in vitro* study that more goat milk casein remained undigested after 30 min of digestion compared to cow milk casein (Inglingstad, Devold et al. 2010). Since most research has been done *in vitro* and results are not conclusive, verification *in vivo* in humans is warranted. Understanding differences in coagulation and gastric emptying of cow and goat milk proteins is of interest because they may influence subsequent serum AA availability.

In summary, *in vitro* results suggest that the differences in digestion between cow and goat milk are due to differences in their casein. Therefore, the current *in vivo* study aimed to quantify gastric digestion and absorption of cow and goat milk-derived casein *in vivo* in humans. We used MRI to evaluate intragastric processes and gastric emptying and examined blood AA concentrations. We hypothesized that goat milk-derived casein has different coagulum characteristics, faster gastric emptying and higher serum AA concentrations compared to cow milk-derived casein.

## METHODS

### Design

The study was a randomized cross-over study in which healthy men underwent gastric MRI scans before and after consumption of 300 ml of a cow milk-derived-casein (cow MC) or goat milk-derived-casein (goat MC) drink. Primary outcomes were gastric emptying (measured by gastric emptying half time (GE-t50) and gastric volume over time) and serum AA concentrations. The secondary outcome was gastric coagulation. Other outcomes were serum glucose, insulin, free fatty acids (FFA) and triglyceride (TG) concentrations and appetite ratings (hunger, fullness, desire to eat, prospective consumption and thirst). The study was conducted according to the principles of the Declaration of Helsinki (October 2013) and registered with the Dutch Trial Register under number NL8137 (accessible through https://trialsearch.who.int/Trial2.aspx?TrialID=NL-OMON28580). All participants signed written informed consent.

#### Sample size

Sample size was calculated based on the primary outcomes, gastric emptying and post prandial AA profile. Given our previous work on gastric emptying of caloric liquids in adults, we know that the gastric emptying half time of 500 ml dairy-based shakes differing in energy density and viscosity (protein content either 6 or 30 g and energy density 0.2 kcal/ml or 1 kcal/ml) has an overall gastric emptying half-time (GE-t50) of 54.7 ± 3.8 min, with the different shakes ranging between 26.5 ± 3.0 min and 81.9 ± 8.3 min. The goat MC drink contains 0.58 kcal/ml. Based on nutrient density the expected SD of the cow MC and the goat MC drinks should be somewhere between 3 and 6. Therefore, we assumed an SD of 6 min for both drinks. In Camps et al. 2016 the average difference in GE-t50 between a 500-ml thin and thick shake was 14.5 (low-calory) and 12.4 min (high-calory). For a 300-ml load the GE-t50 will be smaller. We considered a 4 min difference in gastric emptying half-time between the treatments as the minimum detectable difference. A two-sided test was deemed appropriate, with Zα=1.96, p=0.05 and a power of 90% gives a Zβ of 1.28. Combined with the SD of 6 this leads to the following formula for paired comparisons (Gogtay 2010): n = (1.96 + 1.28)^2 x (6/4)^2 = (3.24)^2 x 1.5= 18 participants.

To ensure sample size was sufficient for the other primary outcome, post prandial AA profile, a second sample size calculation was performed. The estimation for postprandial AA was based on the peak value and the total free AA assessed in the serum after consumption of protein products. For the peak value, a difference of 100 µg/ml was regarded as relevant with an individual difference in peak values of 100 µg/ml (Farnfield, Trenerry et al. 2009, He, Spelbrink et al. 2013). Again we used a two-sided test, with Zα=1.96, p=0.05 and power of 90% gives a Zβ=1.28. This leads to the following formula for paired comparisons: n = (1.96 + 1.28)^2 x (100/100)^2 = (3.24)^2 x 1= 11 participants. Based on the above, we aimed to include 18 participants.

#### Participants

Healthy males were recruited from November 2019 until March 2020 via e-mail using a database of individuals who expressed interest in participating in scientific research. Healthy, non-smoking men with a BMI of 18.5-25 kg/m^2^ were included. Exclusion criteria were cow or goat milk allergy or lactose intolerance (self-reported), gastric disorders or regular gastric complaints, use of proton pump inhibitors or other gastric medication, or a contra-indication to MRI scanning (including, but not limited to pacemakers and defibrillators, intraorbital or intraocular metallic fragments ferromagnetic implants or being claustrophobic). Eighteen healthy men (age 23 ± 1.6 y, BMI 23 ± 1.6 kg/m^2^) participated in the study. The flow diagram can be found in **Supplemental figure 1**.

#### Treatments

Treatments were 300 ml drinks containing 30 g of protein originating from cow micellar casein concentrate powder (FrieslandCampina Refit MCI80) or 30 g goat micellar casein concentrate powder (Ausnutria Dairy Corporation Ltd, pilot plant). The drinks were matched on caloric content, protein, lactose and dry matter content. The amount of the AA leucine was equal. Vanilla extract was added to ensure similarity of taste. The drinks were prepared by slowly reconstituting the caseins at 50°C and thereafter allowing them to fully rehydrate at 4°C overnight using magnetic stirrer. This method assured a more similar viscosity in the drinks (when resolving fast by shaking there was a larger viscosity of the cow MC drink: 200-500 mPa.s and goat MC drink 8-15mPa.s). Three samples of each drink were measured with a rheometer to assess their viscosity using an Anton Paar Physica MCR 301 rheometer equipped with a Couette geometry with a volume of 1 ml. Viscosity was determined at constant shear rate of 100s-1 at 20 °C. The difference in viscosity of the drinks (15.9 ± 3.9 mPa.s for the cow MC drink and 7.2 ± 1.8 mPa.s for the goat MC drink) was deemed not physiologically relevant (for instance milk ‘s viscosity is 2 mPa.s while that of yoghurt is 150 mPa.s which is a much larger difference).

The composition per 300 ml serving of the cow MC drink was 176 kcal, 30 g protein, 90:10 casein to whey ratio, 12 g lactose, 0.77 g fat and 81 g dry matter. For the goat MC drink this was 174 kcal, 30 g protein, 90:10 casein to whey ratio, 12 g lactose, 0.60 g fat, 81 g dry matter. The amino acid profiles of the drinks can be found in **Table 1**. The data were analyzed by Qlip B.V. (lactose, dry matter, fat, protein) and NIZO (casein-to-whey ratio of the goat micellar casein concentrate; for cow micellar casein concentrate the 90:10 casein to whey ratio was stated on the product data sheet).

**Table 1.**
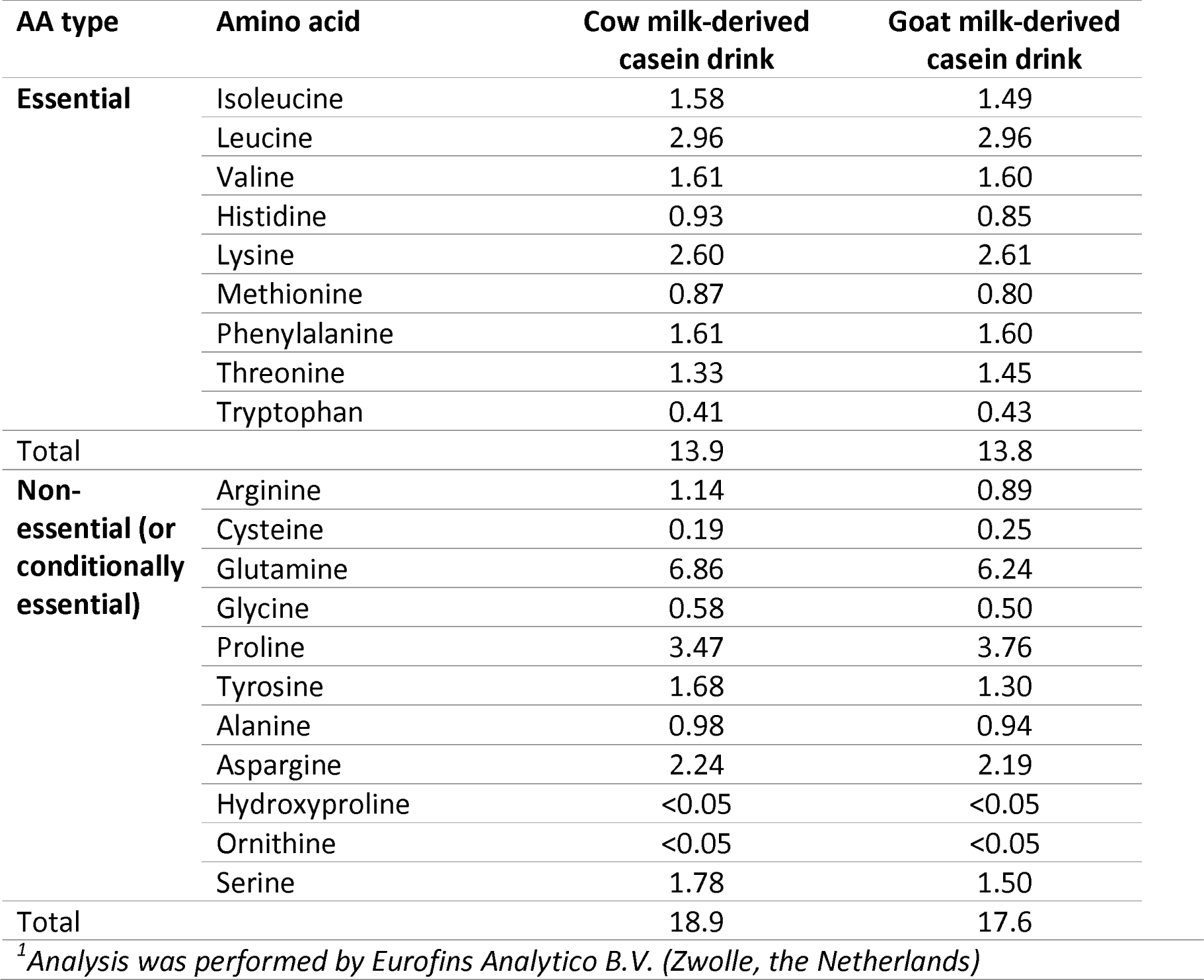
Amino acid profile of cow and goat milk-derived casein drinks (g AA per 300 ml)

| AA type | Amino acid | Cow milk-derived<br>casein drink | Goat milk-derived<br>casein drink |
| --- | --- | --- | --- |
| **Essential** | Isoleucine | 1.58 | 1.49 |
|  | Leucine | 2.96 | 2.96 |
|  | Valine | 1.61 | 1.60 |
|  | Histidine | 0.93 | 0.85 |
|  | Lysine | 2.60 | 2.61 |
|  | Methionine | 0.87 | 0.80 |
|  | Phenylalanine | 1.61 | 1.60 |
|  | Threonine | 1.33 | 1.45 |
|  | Tryptophan | 0.41 | 0.43 |
| Total |  | 13.9 | 13.8 |
| **Non-essential (or**<br>**conditionally**<br>**essential)** | Arginine | 1.14 | 0.89 |
|  | Cysteine | 0.19 | 0.25 |
|  | Glutamine | 6.86 | 6.24 |
|  | Glycine | 0.58 | 0.50 |
|  | Proline | 3.47 | 3.76 |
|  | Tyrosine | 1.68 | 1.30 |
|  | Alanine | 0.98 | 0.94 |
|  | Asparagine | 2.24 | 2.19 |
|  | Hydroxyproline | <0.05 | <0.05 |
|  | Ornithine | <0.05 | <0.05 |
|  | Serine | 1.78 | 1.50 |
| Total |  | 18.9 | 17.6 |
| <sup>1</sup>Analysis was performed by Eurofins Analytico B.V. (Zwolle, the Netherlands) |  |  |  |

#### Study procedures

Participants arrived after an overnight fast. Eating was allowed until 8 PM on the day before the study. Drinking water was allowed up to one hour before the visit. Participants started their scan session at 8 or at 10 AM and were measured at the same time on both study days. First, a canula was placed in an antecubital vein. Then, participants verbally provided baseline appetite ratings after which the baseline MRI scan was performed and the baseline blood sample was drawn. Subsequently, they consumed one of the 300-ml drinks within two minutes. Participants were randomly allocated by block randomization using https://randomizer.org to receive either the cow or goat MC first. Participants were blind for which drink they received. Gastric MRI scans were performed at baseline and at t = 3, 10, 20, 30, 40, 50 and 60 minutes after the start of ingestion. During the MRI session participants verbally rated hunger, fullness, prospective consumption, thirst and nausea on a scale from 0 (not at all) to a 100 (most imaginable) at each time point (Noble, Clark et al. 2005, Blundell, de Graaf et al. 2010). An overview of a test session is given in **Figure 1**.

**Figure 1.**
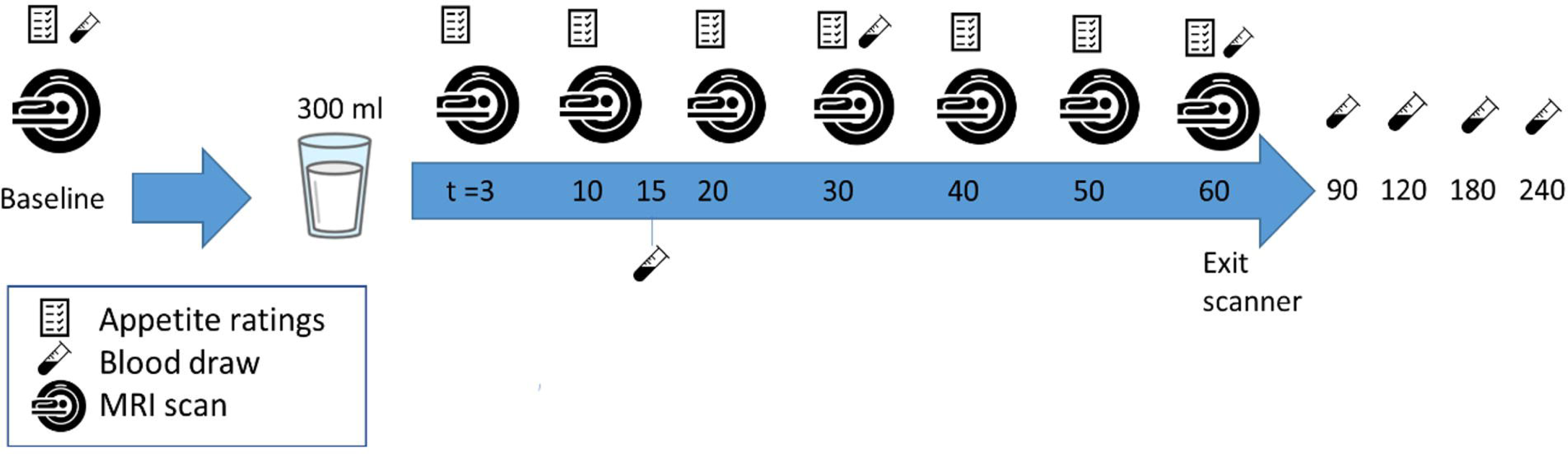
Overview of a test session

#### MRI

Participants were scanned in a supine position with the use of a 3 Tesla Siemens Verio MRI scanner (Siemens AG, Munich, Germany) using a T_2_-weighted spin echo sequence (HASTE, 24 6-mm slices, 2.4 mm gap, 1.19 x 1.19 mm in-plane resolution, TR 850 ms, TE 87 ms, flip angle 112 °C) with breath hold command on expiration to fixate the position of the diaphragm and the stomach. The duration of the scan was approximately 18 seconds. The software Medical Imaging Processing And Visualization (MIPAV, version 11.0.3) (McAuliffe, Lalonde et al. 2001) was used to manually delineate gastric content on each slice and create a corresponding mask image. Gastric content volume on each time point was calculated by multiplying surface area of gastric content per slice with slice thickness including gap distance and summing up the volumes of all slices showing gastric content. In Matlab (version R2023a, multitresh function) a bias field correction was performed using multiplicative intrinsic component optimization (MICO) (Li, Gore et al. 2014). To assess changes in gastric coagulation, image texture analysis of the stomach content was performed using the masks in the software LIFEx (version 7.2.0, Institut national de la santé et de la recherche médicale, France) (Nioche, Orlhac et al. 2018). Homogeneity, coarseness, contrast, and busyness were calculated. These image metrics provide information on the spatial patterns of voxel intensity (Thomas, Qin et al. 2019). The Gray-Level Co-occurrence Matrix (GLCM) method was used for homogeneity (degree of similarity between voxels) and Neighborhood Gray-level Difference Matrix (NGLDM) difference of grey-levels between one voxel and its 26 neighbors in 8 dimensions was used for contrast (local variations), coarseness (spatial rate of change in intensity) and busyness (spatial frequency of changes in intensity). The number of grey-levels for texture metric calculation was set at 64, intensity rescaling at relative (ROI: min/max), and dimension processing at 2D. On each postprandial time point, texture metrics were calculated per slice for the stomach content. Subsequently, a weighted average texture metric was calculated based on the gastric content volume in each slice such that slices with little stomach content contributed less to the average than those with more stomach content. To quantify the (relative) volume of liquid and semi-solid stomach contents the number of lighter (more liquid), intermediate and darker (semi-solid) voxels was calculated by determining two intensity thresholds with the use of Otsu’s method (Otsu 1979) in Matlab, an approach previously used on *in vitro* and *in vivo* MRI images of gastric milk digestion (Mayar, Smeets et al. 2023, van Eijnatten, Camps et al. 2023, Mayar, de Vries et al. 2024). The number of intermediate and darker voxels were summed and interpreted as reflecting voxels in which coagulation took place. This was done because visual inspection of the thresholding results using one threshold showed a poorer separation between lighter and darker gastric contents. In the context of this study, changes in image texture metrics were interpreted as reflecting changes in the degree of coagulation. An example of stomach contents with and without coagulation and the corresponding image texture metrics can be found in **Supplemental figure 2.**

#### Blood parameters

Blood samples were collected at baseline and at t = 15, 30, 60, 90, 120, 180 and 240 min in sodium-fluoride (for glucose), serum (for AA, FFA and insulin) and lithium-heparin (for triglyceride) tubes. After collection, blood in the serum tube was allowed to clot for 30 minutes. Subsequently, all tubes were centrifuged at 1300 g for 10 minutes at 20 °C. After centrifugation, the supernatant was divided into aliquots and stored in a -80°C freezer until analysis. AA, FFA and insulin concentrations were measured at Wageningen University. Glucose and TG concentrations were measured at the clinical chemistry lab of the Gelderse Vallei hospital (Ede, The Netherlands).

AA concentrations were determined using triple quadrupole mass spectrometry (TQMS), with an internal standard and ^13^C reference mix. Glucose concentrations were determined using an Atellica CH Glucose Hexokinase_3 (GluH_3) assay kit and Atellica CH analyzer (Siemens Healthineers, Netherlands). The lower limit of detection (LLOD) was 0.2 mmol/l and intra-assay CVs were at most 4.5%. Serum insulin concentrations were determined using an enzymatic immunoassay kit (ELISA, Mercodia AB, Sweden) with a LLOD of 0.008 mmol/l and intra assay CVs of maximal 6.9%. Serum FFA concentrations were determined using an enzymatic assay kit (Instruchemie, Delfzijl) with a LLOD of 4 mg/dl and intra assay CVs of at m­ost 1.4%. TG concentrations were quantified using an Atellica CH TG enzymatic assay kit and quantified using an Atellica CH analyzer (Siemens Healthineers, Netherlands) with a LLOD of 8 mg/dl and intra assay CVs of at most 1.0%.

#### Statistical analysis

In order to estimate GE-t50, a commonly used summary measure, a curve was fitted to the gastric volume over time of the cow MC and goat MC drink using R statistical software according to the linear-exponential model as developed on the basis of earlier models (Elashoff, Reedy et al. 1982, Fruehauf, Menne et al. 2011). Further analyses were performed in SPSS (version 22, IBM, Armonk, USA). Essential amino acid (EAA), non-essential amino acid (NEAA) and branched-chain amino acid (BCAA) concentrations were calculated by adding individual AA concentrations. GE-t50 was compared between cow MC and goat MC drink with a paired t-test. Normality was confirmed by inspecting QQ plots of the residuals. Overall gastric volume, coagulation (image texture of the gastric contents as reflected in homogeneity, contrast, coarseness and busyness), blood parameters and appetite ratings over time were tested using linear mixed models with treatment, time and treatment*time as fixed factors and baseline values as covariate. Post-hoc t-tests were performed with Tukey’s HSD correction when there were significant effects. In addition, Pearson correlation coefficients were calculated for the associations between image texture metrics at 30 min (chosen because that was the first time point that coagulation was clearly visible) and GE-t50 and blood parameters (4-h AUC of EAA, NEAA, BCAA, glucose and insulin). The significance threshold was set at p = 0.05. Data are expressed as mean ± SEM unless stated otherwise.

## RESULTS

### Gastric emptying

The curves of gastric emptying over time of the cow MC and goat MC drink is shown in **Figure 2**. Gastric emptying half-time was 84.6 ± 23.7 minutes for the cow MC drink, compared to 79.8 ± 24.7 minutes for the goat MC drink (p = 0.395). The mixed model analysis showed no difference in gastric emptying over time between the drinks (MD 0.77, 95% CI [-6.9, 8.5], p = 0.845) and there was no time*treatment interaction effect (p = 0.65). There was a significant effect of time (p < 0.001).

**Figure 2.**
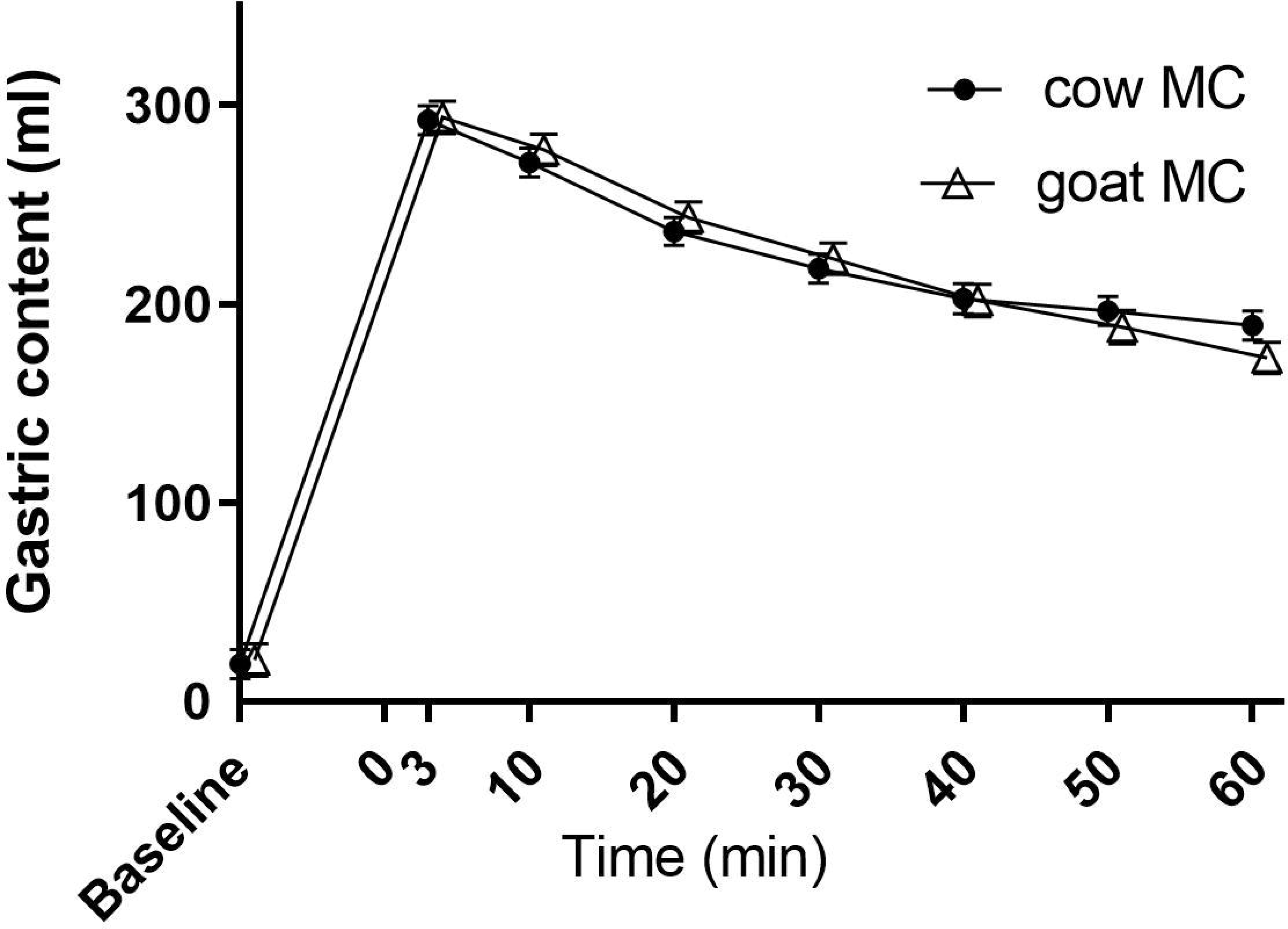
Mean ± SEM gastric content volume over time for the 300 ml cow (cow MC) and goat (goat MC) and milk-derived casein drinks. T=0 min is the start of ingestion. A linear mixed model analysis showed no significant difference between the drinks.

An example of a thresholded stomach image can be found in **Supplemental figure 3**. The percentage of liquid and coagulum volume did not differ between the cow MC and goat MC drink over time (coagulum: MD 0.407 %, 95 % CI [-1.2, 2.0], p = 0.607) and there was no treatment*time interaction effect (p = 0.52). There was a significant decrease in coagulum volume over time (time effect p = 0.002).

### Amino acids

Figures of serum EAA, NEAA and BCAA can be found in Figure 3. Figures of all individual AAs can be found in **Supplemental figure 4 and 5**. Serum concentration of EAA over time was lower for goat MC (MD -110 µmol/L, 95% CI [-162, -58], p < 0.001). Post-hoc t-test showed that this was driven by time points t = 30 (p = 0.002) and t = 180 (p = 0.025) and that there tended to be a lower serum concentration of NEAA for goat MC over time (MD -62.1 µmol/L, 95% CI [-127, 3.4], p = 0.063), driven by time point t = 180 min (p = 0.045). Serum concentration of BCAA was also lower for goat MC (MD -65 µmol/L, 95% CI [-101, -29], p < 0.001). This was driven by time point t = 30 (p = 0.005), and there was a trend at t = 180 (p = 0.051). Time effects where significant for EAA, NEAA and BCAA (p < 0.001). There were no interaction effects.

**Figure 3.**
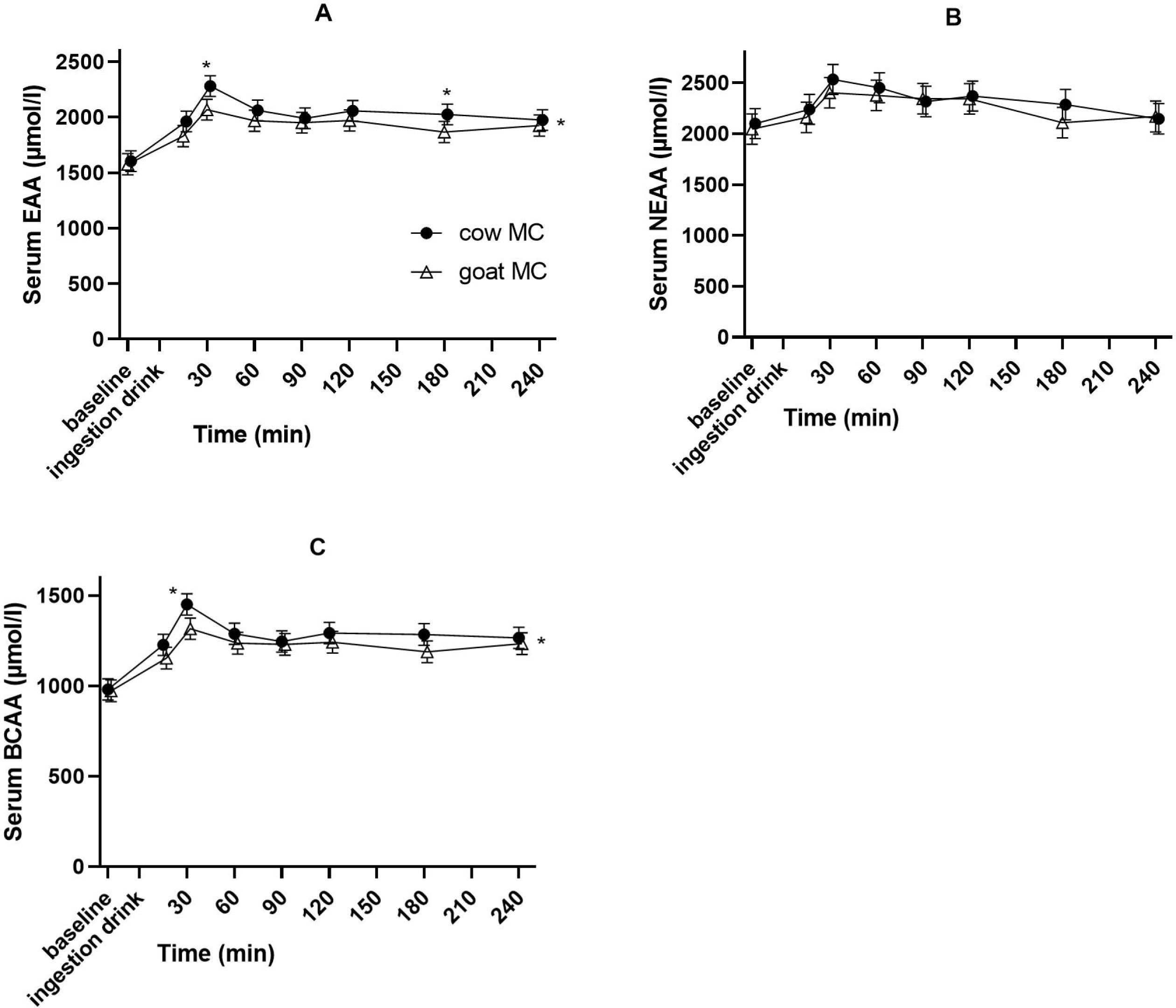
Mean concentration ± SD of (A) serum essential amino acid, (B) non-essential amino acid and (C) branch-chained amino acid concentrations after cow (cow MC) and goat (goat MC) milk-derived casein drink ingestion. *p < 0.05 placed at the right of the graph denotes a significant treatment effect. Above a data point it denotes a significant time point (post-hoc t-test).

### Coagulation

Figure 4 shows examples of MRI images at the level of the stomach showing homogenous stomach content after casein drink consumption and subsequent coagulum formation.

**Figure 4.**
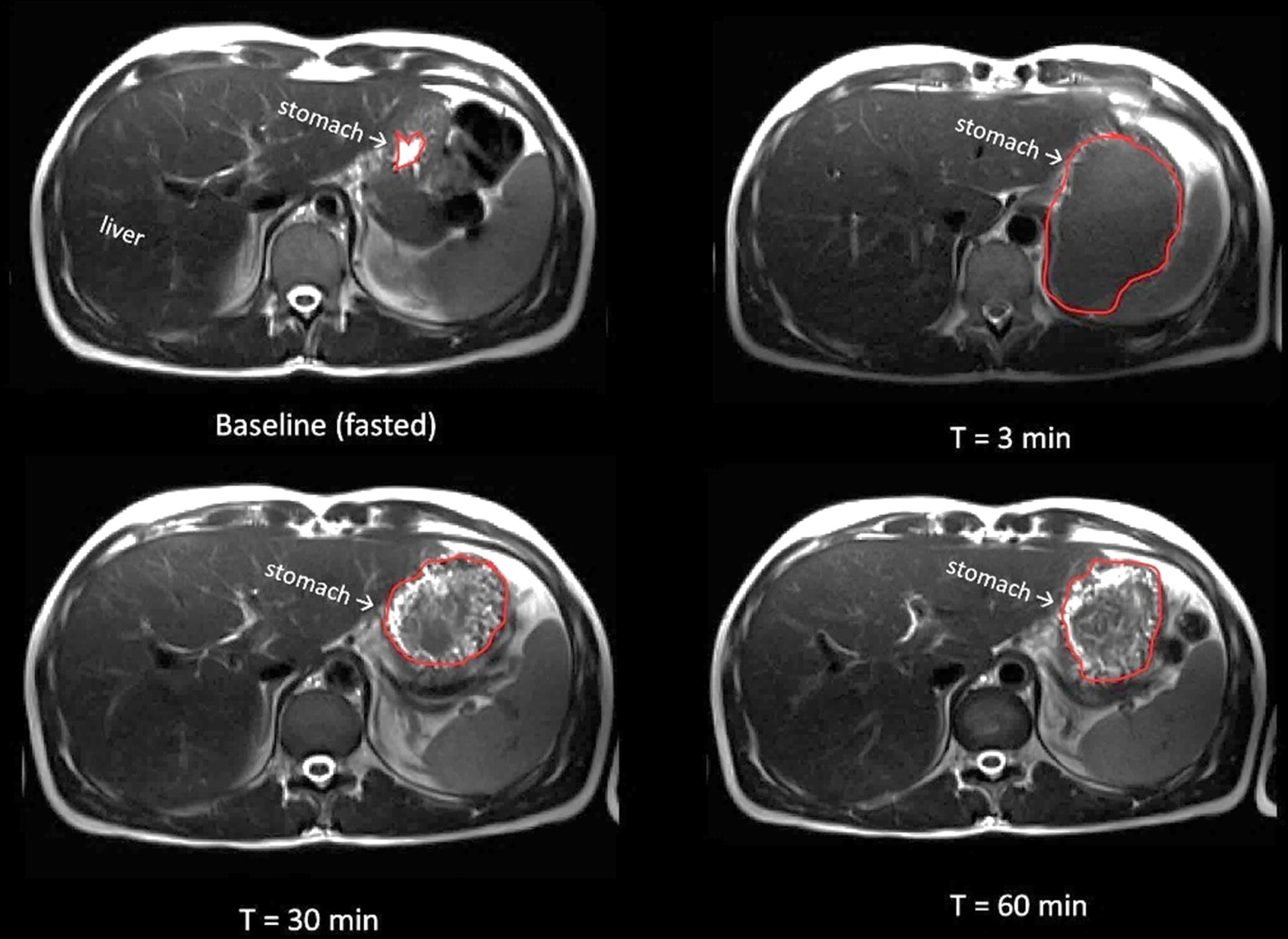
Examples of T_2_-weighted MRI images showing cross-sections through an empty stomach after an overnight fast (baseline) and after 300 ml casein drink consumption. At t = 30 and 60 min coagulation can be observed.

The curves of the image texture metric contrast of the stomach content over time can be found in Figure 5. The other image texture metrics can be found in **Supplemental figure 6**. Contrast was significantly lower for the cow MC than for the goat MC drink (MD 0.010, 95% CI [0.001, 0.020], p = 0.036). Homogeneity (MD -0.003, 95% CI [-0.012, 0.006], p = 0.503), coarseness (MD 0.001, 95% CI [0.000, 0.001], p = 0.310) and busyness (MD -0.008, 95% CI [-0.023, 0.007], p = 0.315) were not significantly different between the drinks. Time effects where significant for all texture metrics (all p < 0.001). There were no interaction effects.

**Figure 5.**
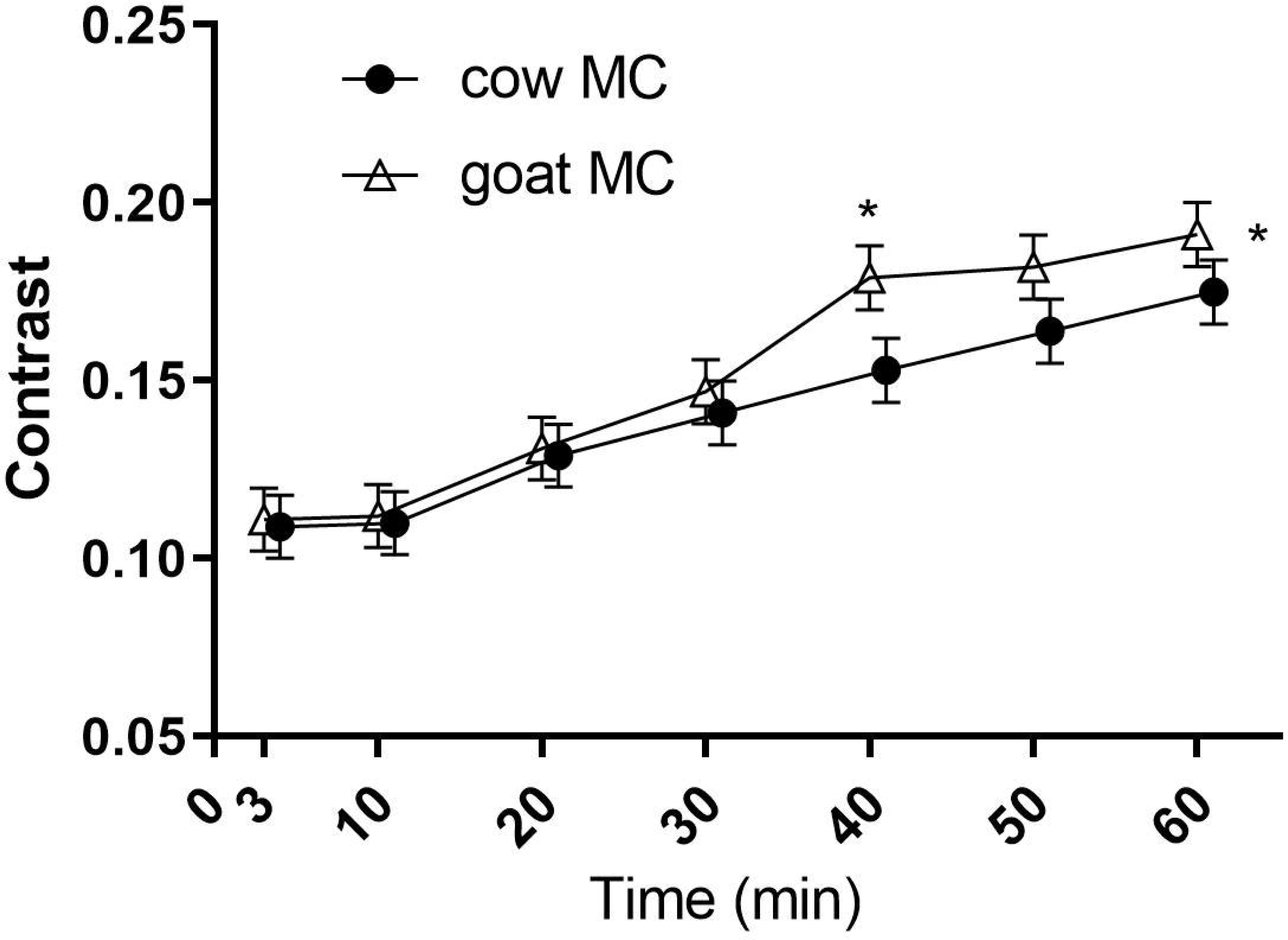
Mean ± SEM of the image texture metric contrast calculated over the stomach content over time after cow (cow MC) and goat (goat MC) milk-derived casein drink ingestion. A linear mixed model analysis showed a significant higher contrast of goat MC (p = 0.036) driven by t = 40 min as seen with a post hoc t-test. A higher contrast reflects a greater degree of structure in the image, which we interpret as a difference in coagulation.

### Glucose and insulin

Overall, glucose concentrations did not differ between the drinks (MD -0.055 mmol/L, 95% CI [-0.14, 0.028], p = 0.19). Only when examining specific timepoints, glucose concentration for the goat MC drink was slightly higher compared to the cow MC drink at t = 30 minutes (MD 0.015 mmol/L, 95% CI [0.42, 0.22], p=0.036). Insulin concentrations did not differ between the cow MC and goat MC drinks (MD -1.66 mIU/L, 95% CI [1.35, -0.16], p = 0.84) (see **Supplemental figure 7**). Time effects where significant for glucose and insulin (p < 0.001). There were no interaction effects.

### Triglycerides and free fatty acids

FFA concentrations were not different between the cow MC and goat MC drinks (MD -0.032 mmol/L, 95% CI [0.034, 0.001], p = 0.948). TG concentrations over time were significantly higher for goat MC (MD 0.074 mmol/L, 95% CI [0.039, 0.109], p = 0.009), even with baseline differences as a covariate in the mixed model analysis. However, there were no differences for any of the time points (see **Supplemental figure 8**). Time effects were significant for FFA and TG (p < 0.001) and there were no interaction effects.

### Appetite ratings

Hunger and thirst did not differ between the treatments (p = 0.61 and 0.29, respectively). Fullness, desire to eat and prospective consumption were significantly lower after goat MC ingestion (p = 0.036, *p* < 0.001, p < 0.001, respectively). However, as can be seen in **Supplemental figure 9**, the differences are small (below 10 units as a mean difference over time).

### Correlations between image texture metrics, blood concentrations and gastric emptying

To explore the associations between image texture metrics as an indicator of the degree of coagulation, blood concentrations and gastric emptying, Pearson correlation coefficients were calculated for image texture metrics at 30 min (because at t=30 min coagulation was visible) and 4-h AUC of blood parameters (EAA, NEAA, BCAA, glucose and insulin). Overall, GE-t50 correlated negatively with contrast (r=-0.413, 95% CI [-0.662, -0.081]) and coarseness (r=-0.38, 95% CI [-0.640, -0.043]). NEAA correlated with insulin (r=0.491, 95% CI [0.152, 0.727]). When divided between treatments: goat MC, EAA, NEAA and BCAA were negatively correlated with coarseness (r=-0.698, 95% CI [-0.887, -0.175], r=- 0.572, 95% CI [-0.837, -0.002] and r = -0.690, 95% CI [-0.891, -0.162] respectively). GE-t50 was positively correlated with insulin (r = 0.558, 95% CI [0.130, 0.790]). For cow MC GE-t50 was positively correlated with image homogeneity and busyness (r = 0.498, 95% CI [0.061, 0.760] and r = 0.568, 95% CI [0.141, 0.787] respectively) and negatively correlated with contrast and coarseness (r = - 0.627, 95% CI [-0.822, - 0.217] and r = - 0.554, 95% CI [-0.780, - 0.134]). All correlations can be found in **Supplemental figure 10**.

## DISCUSSION

This is the first study to explore digestion of cow and goat MC by quantifying gastric emptying, gastric casein coagulation and blood concentrations in humans. Overall, serum concentrations of EAA and BCAA were higher for cow MC. Gastric emptying curves and gastric emptying half-time (GE-t50) were similar for the cow MC and goat MC drinks. The image texture metric contrast was significantly different, suggesting a difference in coagulation. No significant differences between cow MC and goat MC were seen for serum insulin, glucose and FFA concentrations. Serum TG concentrations were significantly higher in goat MC.

Coagulation was visible on the MRI scans for both drinks (see Figure 4). In an attempt to objectively quantify this, image texture metrics were calculated for the stomach contents. A lower homogeneity and higher contrast could reflect ‘more’ coagulation. Even though the actual meaning of differences in image texture metrics in reference to coagulating properties of stomach content requires more research, the difference in the metric ‘contrast’ does support the notion of a difference in coagulation between cow MC and goat MC. However, the metrics homogeneity, coarseness and busyness did not differ between the drinks. It should be taken into account that higher homogeneity could not only reflect a more homogenous liquid, but also the presence of a large and fairly homogenous coagulum. Another aspect to consider is that not only the size, but also the structure is an important characteristic of coagulates. For instance, some coagulates can be firm and have a greater weight and denser structure than less dense coagulates with approximately the same volume (Wang, Ye et al. 2018). Image texture metrics could possibly be used to quantify the density of the coagulum, as MRI can reflect the water content of the coagulum. Notably, MRI image texture parameters are affected by the resolution of the input images and could detect differences in image intensity patterns not appreciable by eye. This requires further validation by concomitant analysis of MRI images and coagulates that differ in size and density. To provide molecular-level information, other MRI techniques are being developed such as measurement of the magnetization transfer ratio and relaxation rates (Deng, Seimys et al. 2022, Deng, Mars et al. 2023, Mayar, Smeets et al. 2023, Mayar, de Vries et al. 2024). These measurements require additional MRI measurements to be recorded, but could be used in follow-up research to examine more subtle differences in protein coagulation *in vivo*.

Cow MC and goat MC coagulates likely differ, since *in vitro* and animal *in vivo* research showed that goat milk coagulum had a softer consistency and less fused protein networks, especially toward a later stage of digestion (Ye, Liu et al. 2019, Roy, Moughan et al. 2022) . We therefore hypothesized that this would lead to a longer gastric retention of coagulum and relatively faster GE for goat milk casein in comparison to cow milk casein. This is seen in recent human *in vivo* work (van Eijnatten, Roelofs et al. 2024). However, we observed that the overall GE was similar and the emptying of coagulum and liquid volume fraction did not differ between cow MC and goat MC. This is in line with recent findings that bovine milk coagulation differences did not affect GE, and GE did not explain differences in AA concentrations (Milan, Barnett et al. 2024).

As expected, even though AA composition of the treatments were largely similar, postprandial serum AA concentrations differed between casein derived from goat and cow milk, which appears to be due in part to differences in their coagulation. Goat milks’ softer and smaller coagulates (Park 2017) would make the proteins more accessible to digestive enzymes (proteases) and lead to more efficient break down of peptide bonds such that goat milk proteins would be faster digested than cow milk proteins (Almaas, Cases et al. 2006). However, our study showed a higher total response of serum AA in the 5 hours after cow milk consumption, which seems contradictive. On the other hand, in an *in vitro* study of Inglingstad et al., who used a two-step digestion model (gastric and duodenal), caseins from goat milk were less digested compared to caseins from cow milk during gastric digestion (Inglingstad, Devold et al. 2010). Interestingly, in our study serum EAA concentrations were higher for cow MC, while we expected higher concentrations for goat MC based on *in vitro* studies (Almaas, Cases et al. 2006, Zhang, Liu et al. 2023). However, we examined caseins in relative isolation where milk fat content was negligible and the mineral profile was different than in the casein micelles present in whole milk. The formation of coagulates interacts with minerals and fat and thus coagulation might be different for a whole food (i.e. in the ‘food matrix’, (Aguilera 2019)). For instance, the interaction of peptides with small fat globules can slow protein breakdown and thereby influence AA availability (Le Feunteun, Barbé et al. 2014, Tunick, Ren et al. 2016). A second factor leading to differences in coagulation might be the difference in buffering capacity between cow and goat milk. Goat milk contains a higher non-protein nitrogen (NPN) amount than cow milk and more NPN contributes to a slower acidification in the stomach (Park 1991). Roy et al also discuss the high degree of variation in casein micelle characteristics within the same species (Roy, Moughan et al. 2022). Within and across species differences in breeds, genetic variants, and phosphorylation sites of the caseins may also add to the variation (Crowley, Kelly et al. 2017). In summary, to our knowledge three factors could have contributed to the discrepancies of our study with other studies: 1) the fact that this is the first human *in vivo* study on gastric digestion, 2) that we assessed casein outside of the food matrix, 3) the buffering capacity and the variation in casein characteristics even within species.

Fullness, desire to eat and prospective consumption were significantly lower for goat MC, but these differences were small. For instance, the mean difference of desire to eat had a mean difference of 7 pts over the 60 measured minutes. A difference below 10% is usually not considered as practically relevant (Flint, Raben et al. 2000). Thus, this finding is in line with the lack of differences in gastric volume over time and coagulum volume over time between the drinks.

We here focused on the casein fractions from cow and goat milk, to be able to study casein coagulation in the absence of interaction effects. Future research should explore casein coagulation within a food matrix, as this could significantly influence protein digestion, involving interactions with components like fat (Cecchinato, Penasa et al. 2012).

In conclusion, cow MC and goat MC show a difference in coagulation as inferred by AA concentrations and supported by image texture analysis *in vivo* in humans. This difference in coagulation did not influence overall gastric emptying or the emptied fraction of the liquid and coagulum volume. Therefore, gastric emptying was not the main driver of AA differences. This warrants further research to examine differences in casein coagulation *in vivo* in the food matrix and how this may affect digestion of milk products, such as infant formula or medical nutrition. This may help to determine the optimal use for cow and goat milk and their protein fractions.

## Data Availability

Deidentified individual participant data that underlie the reported results will be made available upon reasonable request.

## ACKNOWLEDGEMENTS

EE, GC, WR and PS designed the research; EE conducted the research. EE analysed the data and drafted the paper. GC, WR, LP and PS revised the manuscript critically for important intellectual content. PS had primary responsibility for final content. All authors read and approved the final manuscript. This study was funded by Ausnutria Dairy Corporation Ltd and WR and LP were employed by Ausnutria Dairy Corporation Ltd. All other authors declare no further conflict of interest. We thank Julia Roelofs, Mireille Schipper, Jip Jordaan and Lisa van den Berg for assisting with data collection and Jacques Vervoort and Sebas Wesseling for the amino acid analysis. The use of the 3T MRI was made possible by WUR Shared Research Facilities.

## Disclosures

Elise J.M. van Eijnatten: no conflicts of interest

Guido Camps: no conflicts of interest

Wolf Rombouts: during the study employed as a scientist at Ausnutria Dairy Corporation Ltd

Linette Pellis: during the study employed as a scientist at Ausnutria Dairy Corporation Ltd

Paul A.M. Smeets: no conflicts of interest

## Author contributions to manuscript

WR, GC and PS designed the research; EE conducted the research. EE analysed the data and drafted the paper. WR and LP helped interpreting results. GC, WR and PS revised the manuscript critically for important intellectual content. PS had primary responsibility for final content. All authors read and approved the final manuscript.

Data Transparency Statement: deidentified individual participant data that underlie the reported results will be made available upon reasonable request.

## List of abbreviations

95% CI: 95% confidence interval
AA: Amino acids
AUC: Area under the curve
BCAA: Branch chained amino acids
BMI: Body mass index
Cow MC: Cow milk-derived micellar casein
EAA: Essential amino acids
FFA: Free fatty acids
GE: Gastric emptying
GE-t50: Gastric emptying half time
Goat MC: Goat milk-derived micellar casein
MD: Mean difference
MRI: Magnetic resonance imaging
NEAA: Non-essential amino acids
TG: Triglycerides
UHT: Ultra-high temperature processing

## Notes

### Clinical Trial

NL8137

### Author Declarations

Ethics committee of Wageningen University gave ethical approval for this work.

### Summary of Updates

Title, abstract and discussion has been updated.

## REFERENCES

1. Aguilera, J. M. (2019). “The food matrix: implications in processing, nutrition and health.” Critical Reviews in Food Science and Nutrition 59(22): 3612–3629.

2. Almaas, H., A.-L. Cases, T. G. Devold, H. Holm, T. Langsrud, L. Aabakken, T. Aadnoey and G. E. Vegarud (2006). “In vitro digestion of bovine and caprine milk by human gastric and duodenal enzymes.” International Dairy Journal 16(9): 961–968.

3. Atherton, P. J. and K. Smith (2012). “Muscle protein synthesis in response to nutrition and exercise.” The Journal of physiology 590(5): 1049–1057.

4. Blundell, J., C. de Graaf, T. Hulshof, S. Jebb, B. Livingstone, A. Lluch, D. Mela, S. Salah, E. Schuring, H. van der Knaap and M. Westerterp (2010). “Appetite control: methodological aspects of the evaluation of foods.” Obes Rev 11(3): 251–270.

5. Ceballos, L. S., E. R. Morales, G. de la Torre Adarve, J. D. Castro, L. P. Martínez and M. R. S. Sampelayo (2009). “Composition of goat and cow milk produced under similar conditions and analyzed by identical methodology.” Journal of Food Composition and Analysis 22(4): 322–329.

6. Cecchinato, A., M. Penasa, C. Cipolat Gotet, M. De Marchi and G. Bittante (2012). “Short communication: Factors affecting coagulation properties of Mediterranean buffalo milk.” Journal of Dairy Science 95(4): 1709–1713.

7. Claeys, W. L., C. Verraes, S. Cardoen, J. De Block, A. Huyghebaert, K. Raes, K. Dewettinck and L. Herman (2014). “Consumption of raw or heated milk from different species: An evaluation of the nutritional and potential health benefits.” Food Control 42: 188–201.

8. Crowley, S. V., A. L. Kelly, J. A. Lucey and J. A. O’Mahony (2017). “Potential Applications of Non-Bovine Mammalian Milk in Infant Nutrition.” Handbook of Milk of Non-Bovine Mammals: 625–654.

9. Deng, R., M. Mars, A. E. M. Janssen and P. A. M. Smeets (2023). “Gastric digestion of whey protein gels: A randomized cross-over trial with the use of MRI.” Food Hydrocolloids 141: 108689.

10. Deng, R., A. Seimys, M. Mars, A. E. M. Janssen and P. A. M. Smeets (2022). “Monitoring pH and whey protein digestion by TD-NMR and MRI in a novel semi-dynamic in vitro gastric simulator (MR-GAS).” Food Hydrocolloids 125: 107393.

11. Do, Q. N., M. A. Lewis, A. J. Madhuranthakam, Y. Xi, A. A. Bailey, R. E. Lenkinski and D. M. Twickler (2019). “Texture analysis of magnetic resonance images of the human placenta throughout gestation: A feasibility study.” PloS one 14(1): e0211060–e0211060.

12. Eijnatten, E. J. M. v., J. J. M. Roelofs, G. Camps, T. Huppertz, T. T. Lambers and P. A. M. Smeets (2023). “Gastric coagulation and postprandial amino acid absorption of milk is affected by mineral composition: a randomized crossover trial.” medRxiv: 2023.2009.2013.23295475.

13. Elashoff, J. D., T. J. Reedy and J. H. Meyer (1982). “Analysis of gastric emptying data.” Gastroenterology 83(6): 1306–1312.

14. Fardet, A., D. Dupont, L.-E. Rioux and S. L. Turgeon (2019). “Influence of food structure on dairy protein, lipid and calcium bioavailability: A narrative review of evidence.” Critical Reviews in Food Science and Nutrition 59(13): 1987–2010.

15. Farnfield, M. M., C. Trenerry, K. A. Carey and D. Cameron-Smith (2009). “Plasma amino acid response after ingestion of different whey protein fractions.” International Journal of Food Sciences and Nutrition 60(6): 476–486.

16. Flint, A., A. Raben, J. E. Blundell and A. Astrup (2000). “Reproducibility, power and validity of visual analogue scales in assessment of appetite sensations in single test meal studies.” International Journal of Obesity 24(1): 38–48.

17. Fruehauf, H., D. Menne, M. A. Kwiatek, Z. Forras-Kaufman, E. Kaufman, O. Goetze, M. Fried, W. Schwizer and M. Fox (2011). “Inter-observer reproducibility and analysis of gastric volume measurements and gastric emptying assessed with magnetic resonance imaging.” Neurogastroenterol Motil 23(9): 854–861.

18. Gogtay, N. J. (2010). “Principles of sample size calculation.” Indian J Ophthalmol 58(6): 517–518.

19. He, T., R. E. J. Spelbrink, B. J. Witteman and M. L. F. Giuseppin (2013). “Digestion kinetics of potato protein isolates in vitro and in vivo.” International Journal of Food Sciences and Nutrition 64(7): 787–793.

20. Hettinga, K., L. Pellis, W. Rombouts, X. Du, G. Grigorean and B. Lönnerdal (2023). “Effect of pH and protein composition on proteolysis of goat milk proteins by pepsin and pancreatin.” Food Research International 173: 113294.

21. Hodgkinson, A. J., O. A. M. Wallace, I. Boggs, M. Broadhurst and C. G. Prosser (2018). “Gastric digestion of cow and goat milk: Impact of infant and young child in vitro digestion conditions.” Food chemistry 245: 275–281.

22. Huppertz, T. and L. W. Chia (2021). “Milk protein coagulation under gastric conditions: A review.” International Dairy Journal 113: 104882.

23. Ingham, B., A. Smialowska, N. M. Kirby, C. Wang and A. J. Carr (2018). “A structural comparison of casein micelles in cow, goat and sheep milk using X-ray scattering.” Soft Matter 14(17): 3336–3343.

24. Inglingstad, R. A., T. G. Devold, E. K. Eriksen, H. Holm, M. Jacobsen, K. H. Liland, E. O. Rukke and G. E. Vegarud (2010). “Comparison of the digestion of caseins and whey proteins in equine, bovine, caprine and human milks by human gastrointestinal enzymes.” Dairy science & technology 90(5): 549–563.

25. Lagrange, V., D. Whitsett and C. Burris (2015). “Global Market for Dairy Proteins.” Journal of Food Science 80(S1): A16–A22.

26. Le Feunteun, S., F. Barbé, D. Rémond, O. Ménard, Y. Le Gouar, D. Dupont and B. Laroche (2014). “Impact of the Dairy Matrix Structure on Milk Protein Digestion Kinetics: Mechanistic Modelling Based on Mini-pig In Vivo Data.” Food and Bioprocess Technology 7(4): 1099–1113.

27. Li, C., J. C. Gore and C. Davatzikos (2014). “Multiplicative intrinsic component optimization (MICO) for MRI bias field estimation and tissue segmentation.” Magn Reson Imaging 32(7): 913–923.

28. Maathuis, A., R. Havenaar, T. He and S. Bellmann (2017). “Protein Digestion and Quality of Goat and Cow Milk Infant Formula and Human Milk Under Simulated Infant Conditions.” J Pediatr Gastroenterol Nutr 65(6): 661–666.

29. Mahé, S., N. Roos, R. Benamouzig, L. Davin, C. Luengo, L. Gagnon, N. Gaussergès, J. Rautureau and D. Tomé (1996). “Gastrojejunal kinetics and the digestion of [15N]beta-lactoglobulin and casein in humans: the influence of the nature and quantity of the protein.” The American Journal of Clinical Nutrition 63(4): 546–552.

30. Mayar, M., M. de Vries, P. Smeets, J. van Duynhoven and C. Terenzi (2024). “MRI assessment of pH and coagulation during semi-dynamic in vitro gastric digestion of milk proteins.” Food Hydrocolloids 152: 109866.

31. Mayar, M., P. Smeets, J. van Duynhoven and C. Terenzi (2023). “In vitro 1H MT and CEST MRI mapping of gastro-intestinal milk protein breakdown.” Food Structure 36: 100314.

32. McAuliffe, M. J., F. M. Lalonde, D. McGarry, W. Gandler, K. Csaky and B. L. Trus (2001). Medical Image Processing, Analysis and Visualization in clinical research. Proceedings 14th IEEE Symposium on Computer-Based Medical Systems. CBMS 2001.

33. Milan, A. M., M. P. G. Barnett, W. C. McNabb, N. C. Roy, S. Coutinho, C. L. Hoad, L. Marciani, S. Nivins, H. Sharif, S. Calder, P. Du, A. A. Gharibans, G. O’Grady, K. Fraser, D. Bernstein, S. M. Rosanowski, P. Sharma, A. Shrestha and R. F. Mithen (2024). “The impact of heat treatment of bovine milk on gastric emptying and nutrient appearance in peripheral circulation in healthy females: a randomized controlled trial comparing pasteurized and ultra-high temperature milk.” Am J Clin Nutr.

34. Miller, B. A. and C. D. Lu (2019). “Current status of global dairy goat production: an overview.” Asian-Australas J Anim Sci 32(8): 1219–1232.

35. Mulet-Cabero, A.-I., A. R. Mackie, P. J. Wilde, M. A. Fenelon and A. Brodkorb (2019). “Structural mechanism and kinetics of in vitro gastric digestion are affected by process-induced changes in bovine milk.” Food hydrocolloids 86: 172–183.

36. Nioche, C., F. Orlhac, S. Boughdad, S. Reuzé, J. Goya-Outi, C. Robert, C. Pellot-Barakat, M. Soussan, F. Frouin and I. Buvat (2018). “LIFEx: A Freeware for Radiomic Feature Calculation in Multimodality Imaging to Accelerate Advances in the Characterization of Tumor Heterogeneity.” Cancer Research 78(16): 4786.

37. Noble, B., D. Clark, M. Meldrum, H. Ten Have, J. Seymour, M. Winslow and S. Paz (2005). “The measurement of pain, 1945-2000.” Journal of Pain and Symptom Management 29(1): 14–21.

38. Otsu, N. (1979). “A Threshold Selection Method from Gray-Level Histograms.” IEEE Transactions on Systems, Man, and Cybernetics 9(1): 62–66.

39. Park, Y. W. (1991). “Relative Buffering Capacity of Goat Milk, Cow Milk, Soy-Based Infant Formulas, and Commercial Nonprescription Antacid Drugs1, 2.” Journal of Dairy Science 74(10): 3326–3333.

40. Park, Y. W. (2017). Goat Milk – Chemistry and Nutrition. Handbook of Milk of Non-Bovine Mammals: 42–83.

41. Roy, D., P. J. Moughan, A. Ye, S. M. Hodgkinson, N. Stroebinger, S. Li, A. C. Dave, C. A. Montoya and H. Singh (2022). “Structural changes during gastric digestion in piglets of milk from different species.” Journal of Dairy Science.

42. Thomas, R., L. Qin, F. Alessandrino, S. P. Sahu, P. J. Guerra, K. M. Krajewski and A. Shinagare (2019). “A review of the principles of texture analysis and its role in imaging of genitourinary neoplasms.” Abdom Radiol (NY) 44(7): 2501–2510.

43. Trommelen, J., M. E. G. Weijzen, J. van Kranenburg, R. A. Ganzevles, M. Beelen, L. B. Verdijk and L. J. C. van Loon (2020). “Casein Protein Processing Strongly Modulates Post-Prandial Plasma Amino Acid Responses In Vivo in Humans.” Nutrients 12(8).

44. Tunick, M. H., D. X. Ren, D. L. Van Hekken, L. Bonnaillie, M. Paul, R. Kwoczak and P. M. Tomasula (2016). “Effect of heat and homogenization on in vitro digestion of milk.” Journal of Dairy Science 99(6): 4124–4139.

45. van Eijnatten, E. J. M., G. Camps, M. Guerville, V. Fogliano, K. Hettinga and P. A. M. Smeets (2023). “Milk coagulation and gastric emptying in women experiencing gastrointestinal symptoms after ingestion of cow’s milk.” Neurogastroenterology & Motility **n/a**(n/a): e14696.

46. van Eijnatten, E. J. M., J. J. M. Roelofs, G. Camps, T. Huppertz, T. T. Lambers and P. A. M. Smeets (2024). “Gastric coagulation and postprandial amino acid absorption of milk is affected by mineral composition: a randomized crossover trial.” Food Funct 15(6): 3098–3107.

47. van Vliet, S., N. A. Burd and L. J. van Loon (2015). “The Skeletal Muscle Anabolic Response to Plant-versus Animal-Based Protein Consumption.” The Journal of Nutrition 145(9): 1981–1991.

48. Wang, X., A. Ye, Q. Lin, J. Han and H. Singh (2018). “Gastric digestion of milk protein ingredients: Study using an in vitro dynamic model.” Journal of Dairy Science 101(8): 6842–6852.

49. Ye, A., J. Cui, E. Carpenter, C. Prosser and H. Singh (2019). “Dynamic in vitro gastric digestion of infant formulae made with goat milk and cow milk: Influence of protein composition.” International Dairy Journal 97: 76–85.

50. Ye, A., W. Liu, J. Cui, X. Kong, D. Roy, Y. Kong, J. Han and H. Singh (2019). “Coagulation behaviour of milk under gastric digestion: Effect of pasteurization and ultra-high temperature treatment.” Food Chemistry 286: 216–225.

51. Zhang, J., D. Liu, Y. Xie, J. Yuan, K. Wang, X. Tao, Y. Hemar, J. M. Regenstein, X. Liu and P. Zhou (2023). “Gastrointestinal digestibility of micellar casein dispersions: Effects of caprine vs bovine origin, and partial colloidal calcium depletion using in vitro digestion models for the adults and elderly.” Food Chemistry 416: 135865.

