## Supplemental information for "Gastric digestion and changes in serum amino acid concentrations after consumption of casein from cow and goat milk: a randomized crossover trial in healthy men"

**Supplementary information**

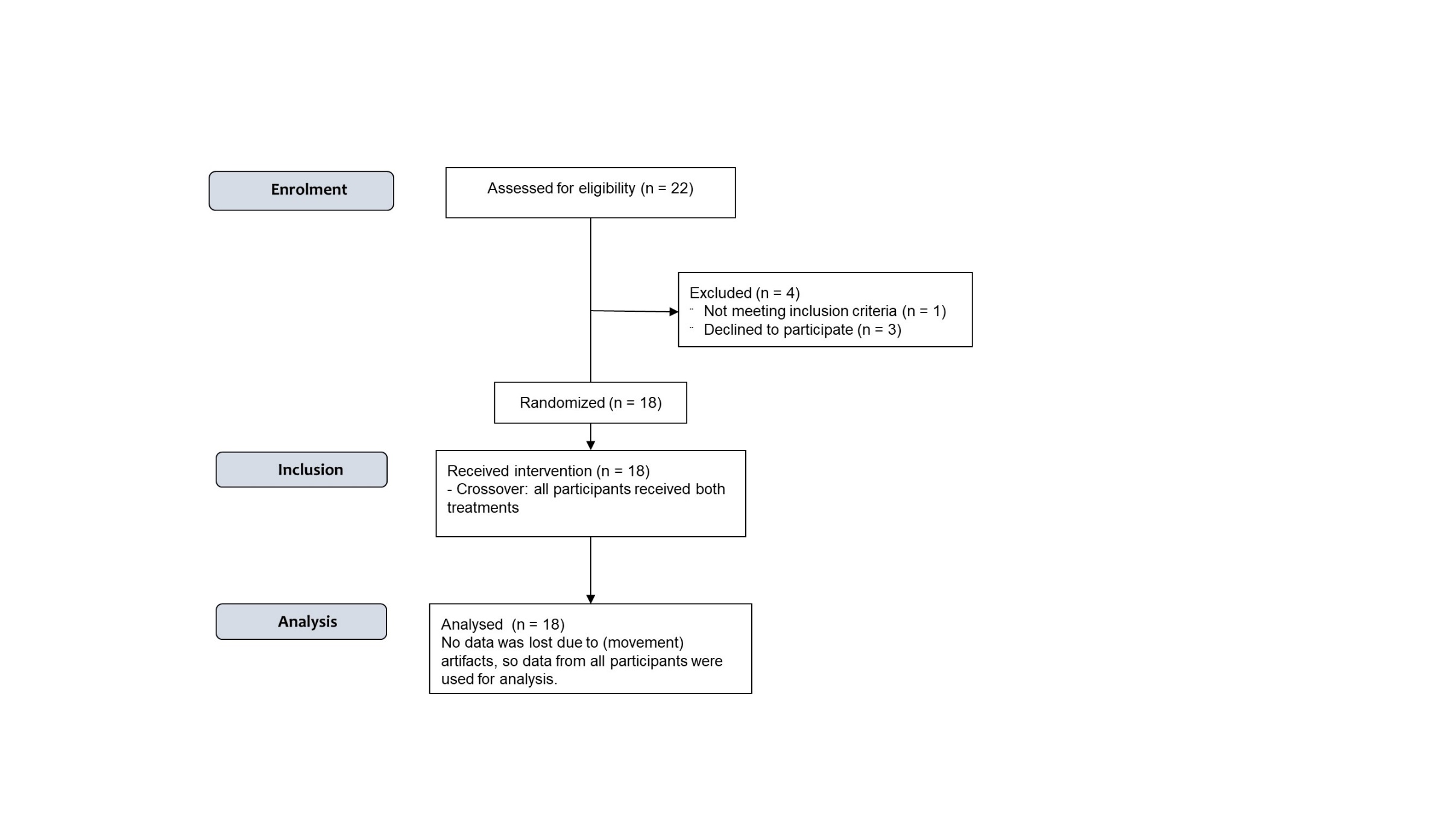

**Supplemental figure 1.** Study flow

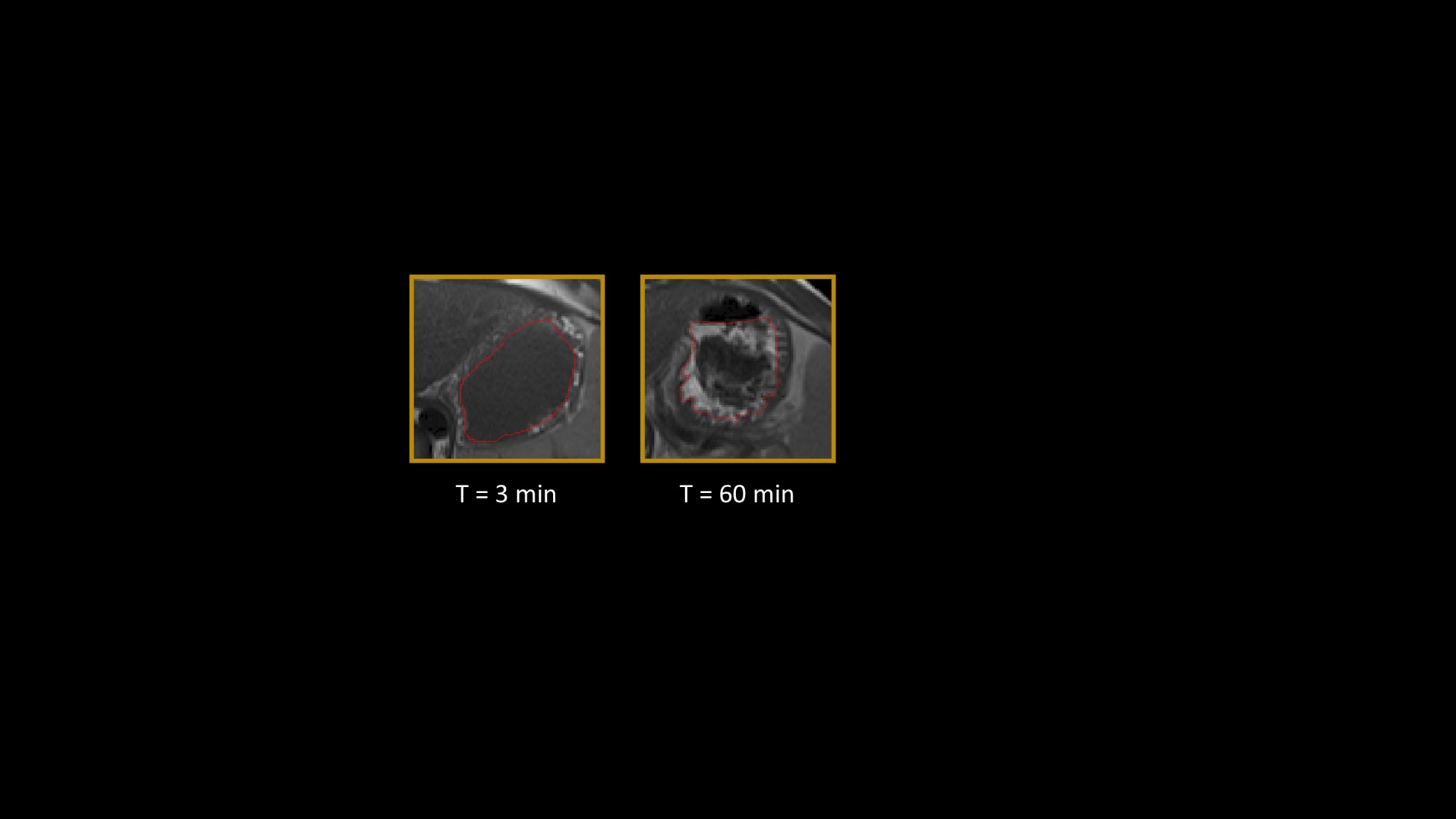

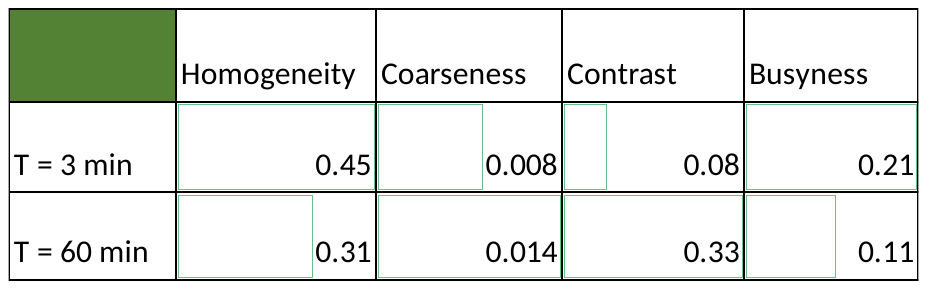

**Supplemental figure 2.** An example of stomach MRI cross-sections with and without coagulation and their corresponding image texture measures for the whole stomach contents.

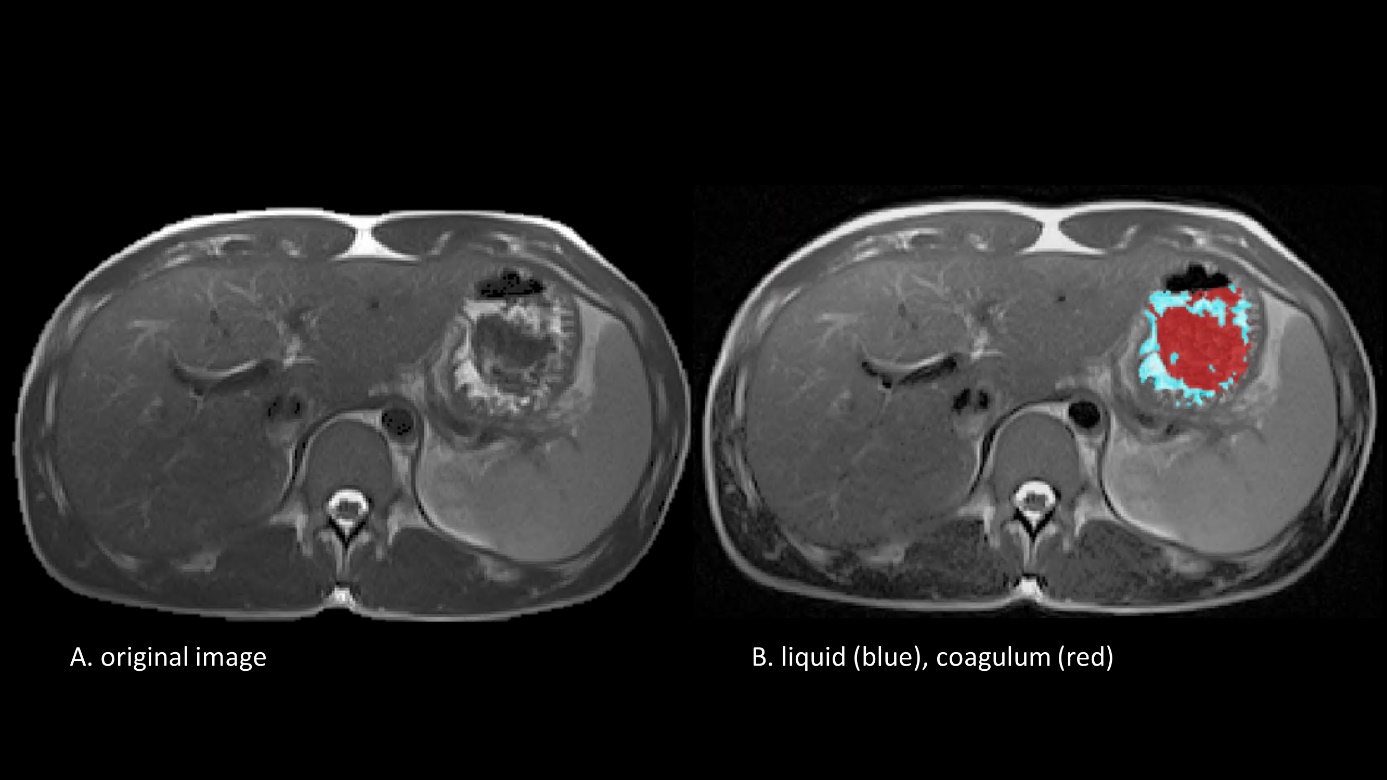

**Supplemental figure 3.** Examples of abdominal MRI images showing cross-sections through a stomach at T = 60 min with A showing the original image and B the same image with color-coded stomach content obtained by thresholding: liquid is shown as blue and semi-solid as red.

**Supplemental table 1.** *Components of goat and cow milk-derived casein drinks per 300 ml*

| Component | Goat milk-derived protein drink | Cow milk-derived protein drink |
| --- | --- | --- |
| Goat MCC^1^ powder (g) | 49.2 | - |
| Cow MCC^1^ powder (g) | - | 38.1 |
| Cow UF^2^ permeate powder (g) | - | 10.1 |
| Water (g) | 248 | 249 |
| Vanilla powder (g) | 3.00 | 3.00 |
| Total (g) | 300.2 | 300.2 |
| *^1^Micellar casein concentrate*  *^2^Ultrafiltration* | | |

| 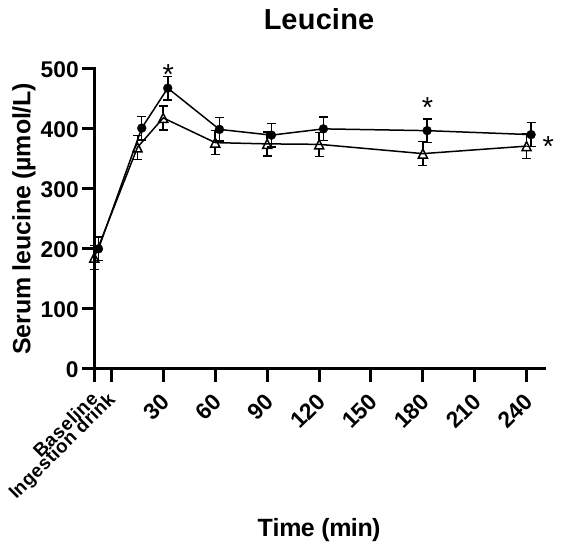 | 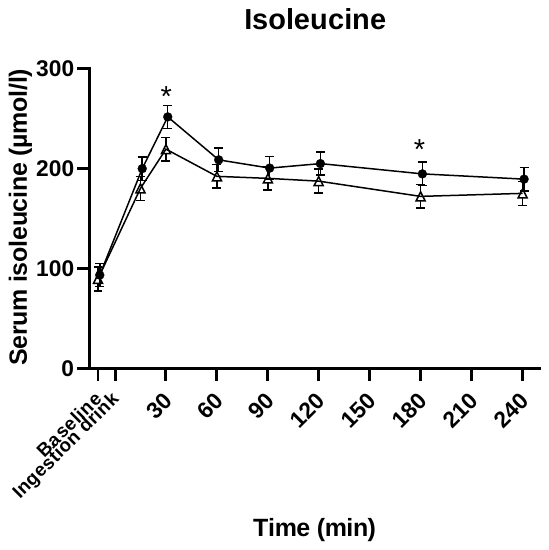 |
| --- | --- |
| 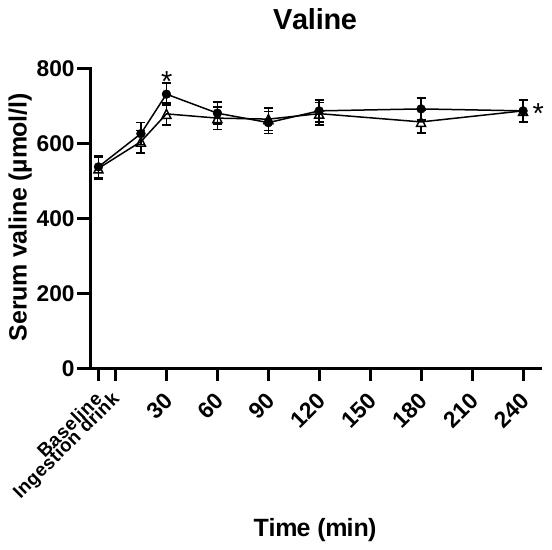 | 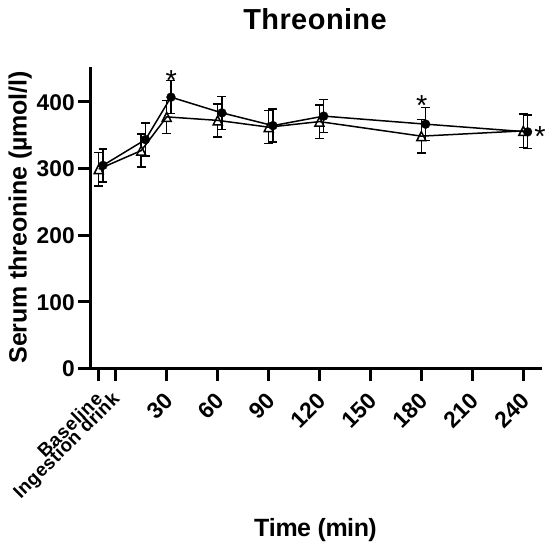 |
| 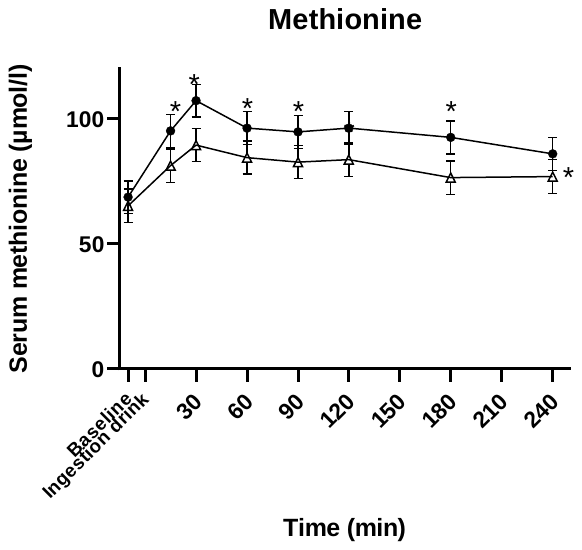 | 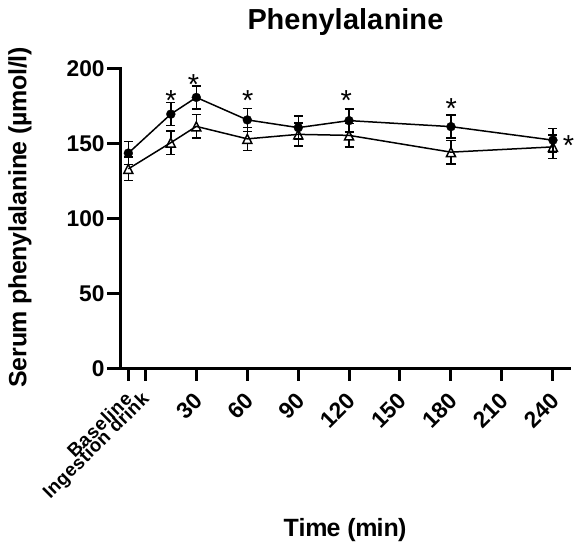 |
| 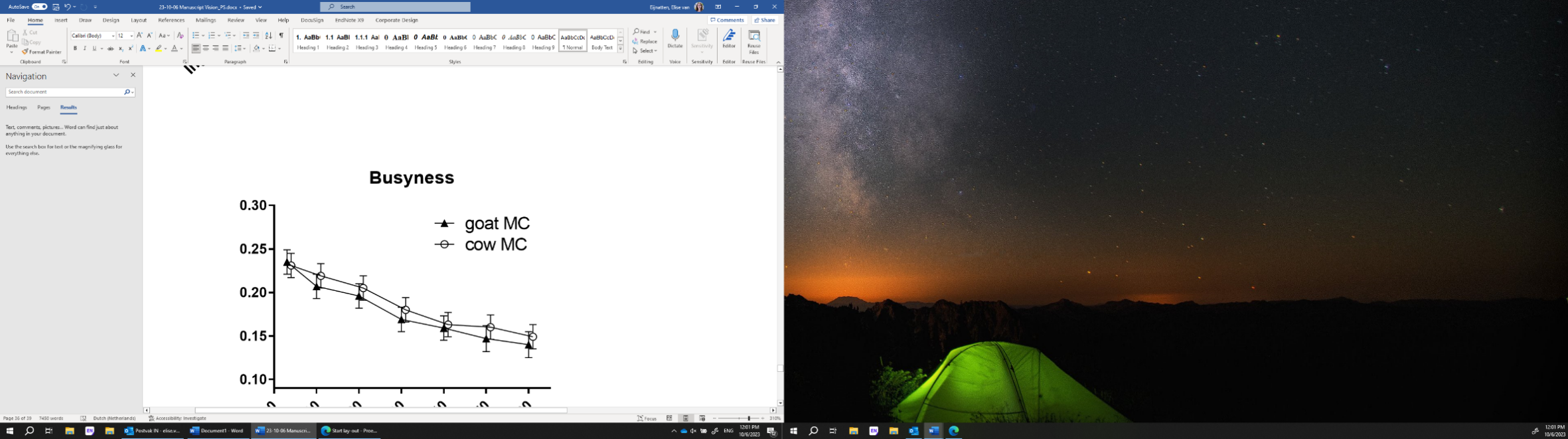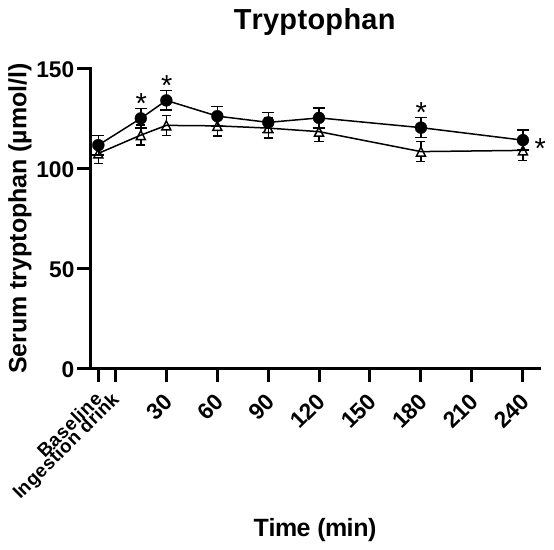 | |

**Supplemental figure 4.** *Mean ± SEM serum concentrations of essential amino acids for cow and goat milk-derived casein drinks. *placed at the right of the graph denotes a significant treatment effect (p<0.05). Above a data point it denotes a significant time point (post-hoc t-test p<0.05).*

| 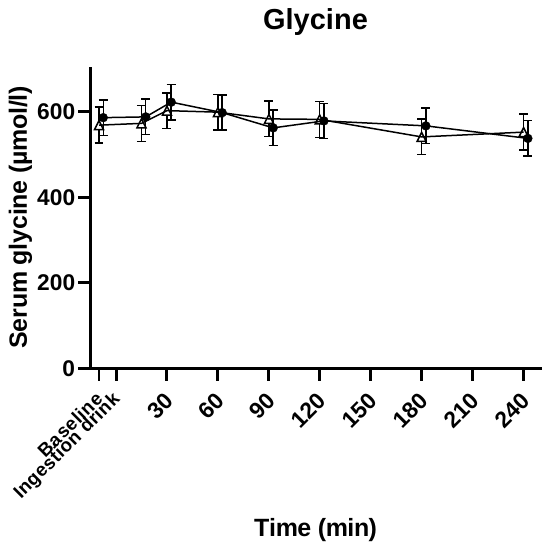 | 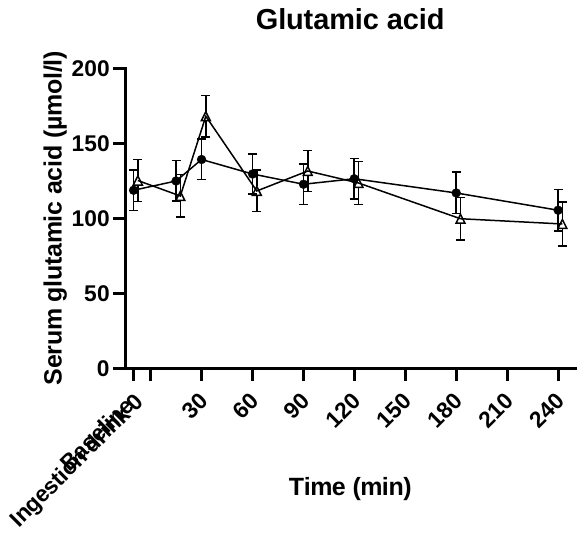 |
| --- | --- |
| 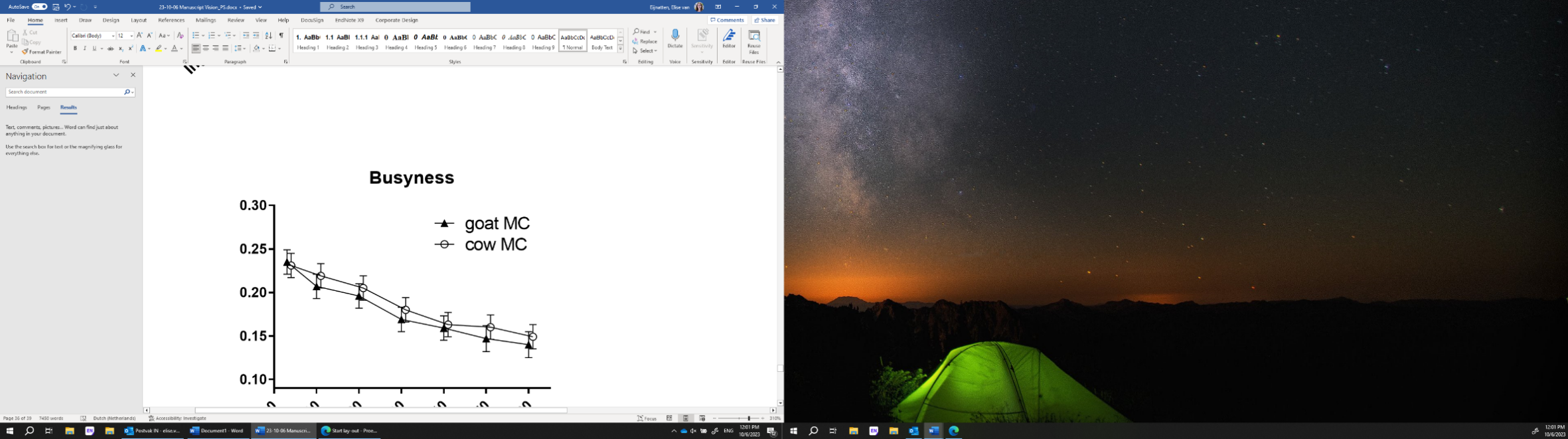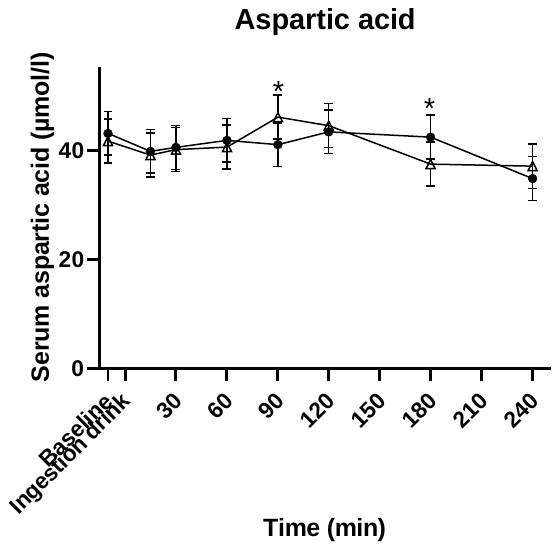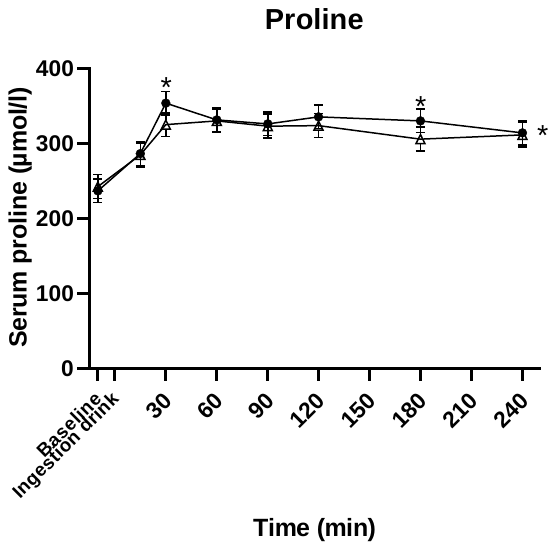 | 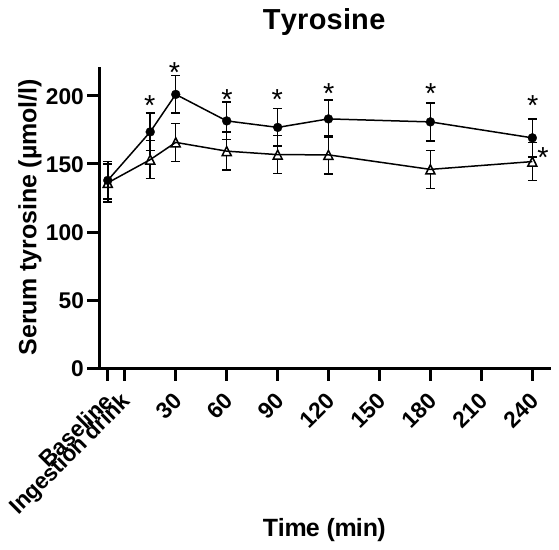 |
| 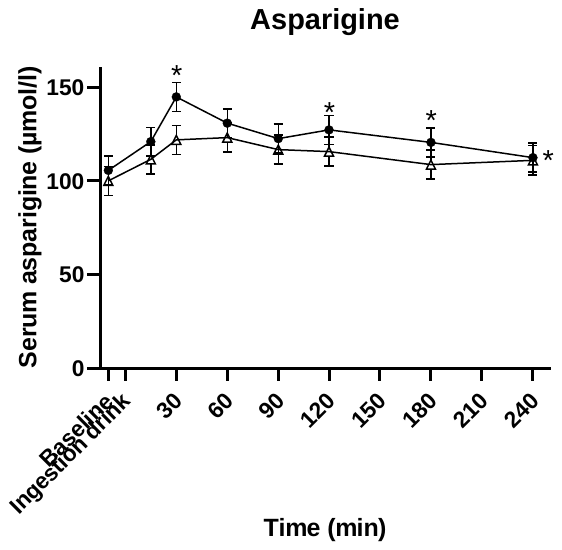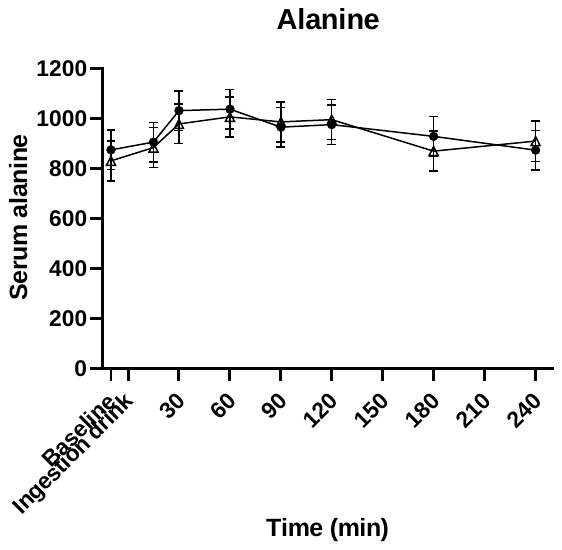 | |

**Supplemental figure 5.** *Mean* ± *SEM serum concentrations of non-essential amino acids* *for cow and goat casein drinks displayed as relative concentrations over time (mean* ± *SEM). * placed at the right of the graph denotes a significant treatment effect (p<0.05). Above a data point it denotes a significant time point (post-hoc test p<0.05).*

1

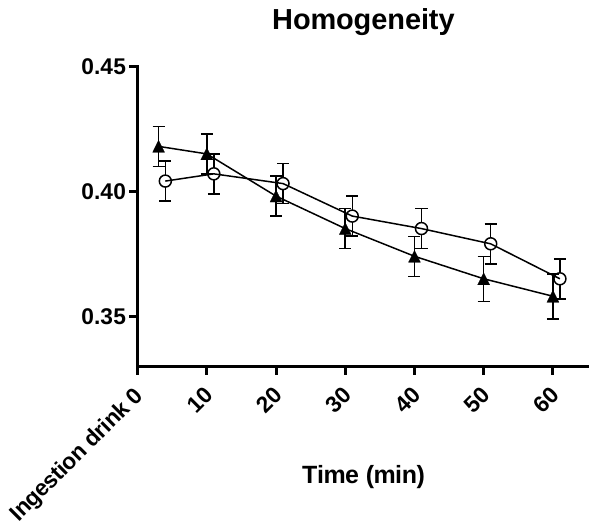

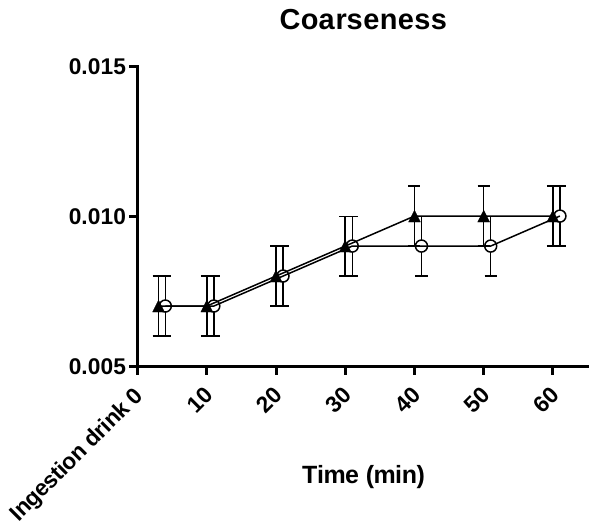

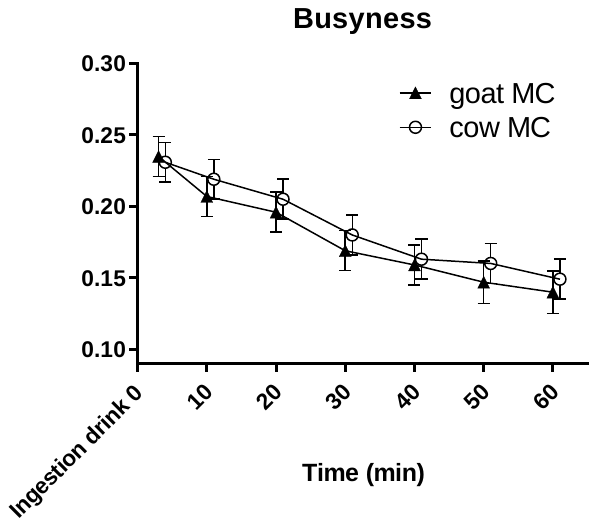

**Supplementary figure 6.** *Mean* ± *SEM of image texture measures homogeneity, coarseness, contrast and busyness of stomach content for cow and goat milk-derived casein drinks over time (a.u.). A higher image contrast reflects a greater degree of structure (possibly coagulation).*

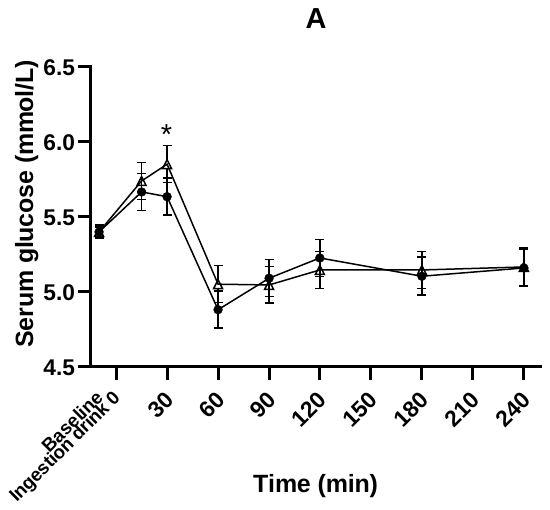

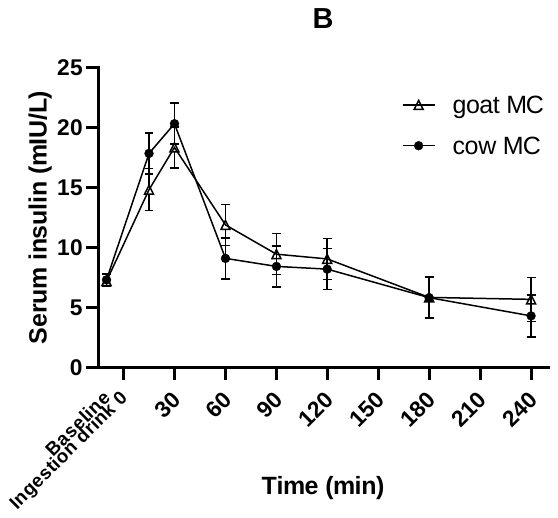

**Supplemental figure 7.** *Mean ± SEM concentrations of glucose (A) and insulin (B) over time after cow and goat milk-derived casein drink ingestion (mmol/L and mIU/L) over time. *placed at the right of the graph denotes a significant treatment effect (p<0.05), above a data point it denotes a significant time point (post-hoc test p<0.05).*

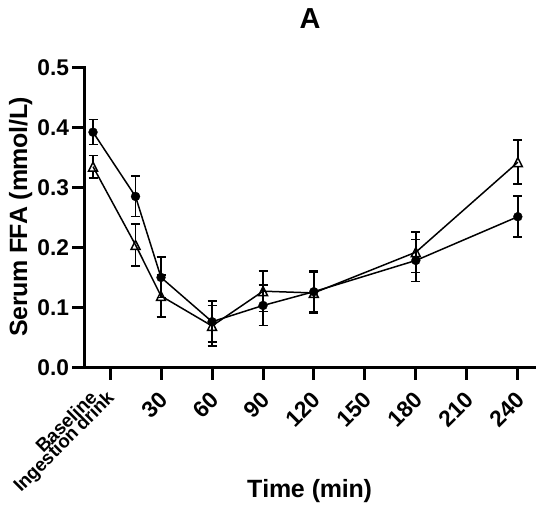

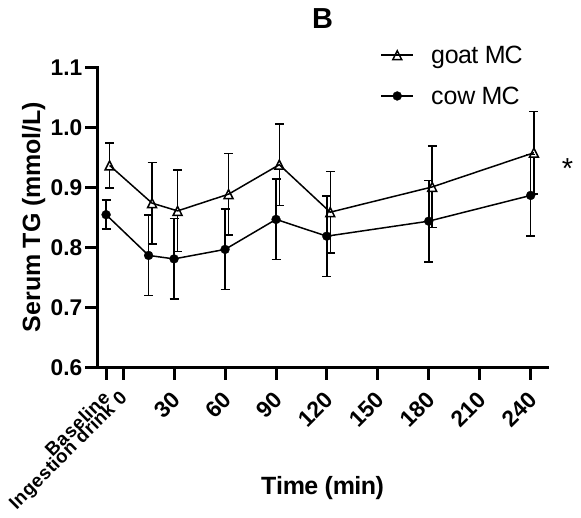

**Supplemental figure 8.** *Mean* ± *SEM serum concentrations of free fatty acids (A) and triglycerides (B) for cow and goat milk-derived casein drink ingestion displayed as mmol/L over time. *placed at the right of the graph it denotes a significant treatment effect (p<0.05). Above a data point it denotes a significant time point (post-hoc test p<0.05).*

| 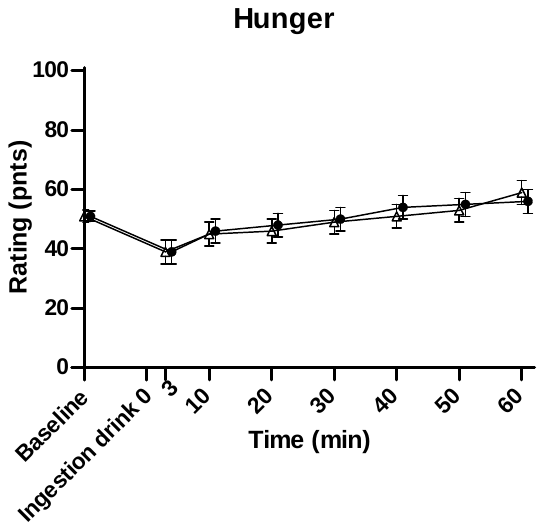 | 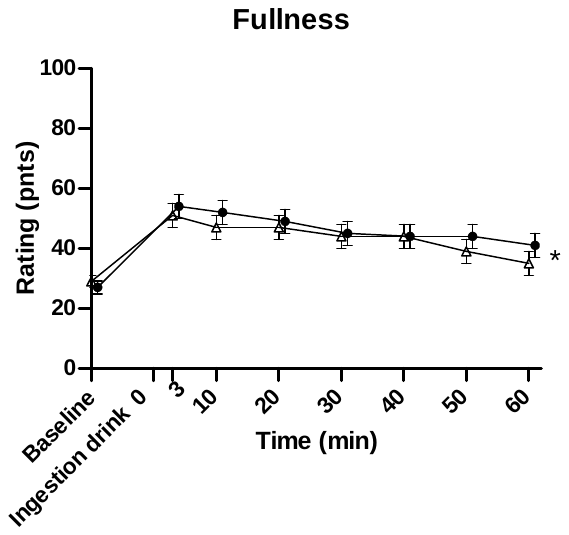 |
| --- | --- |
| 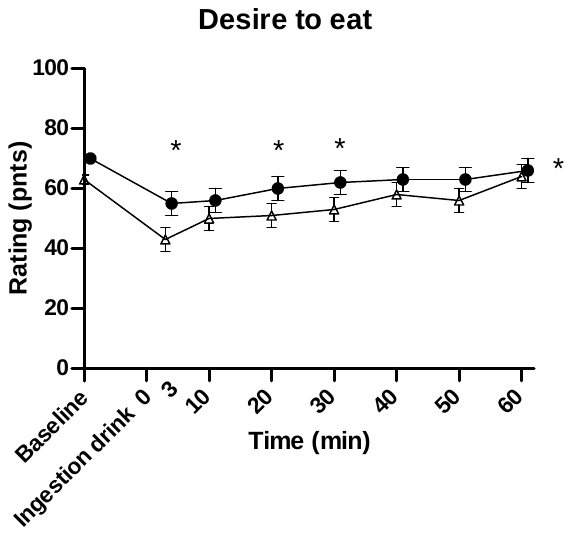 | 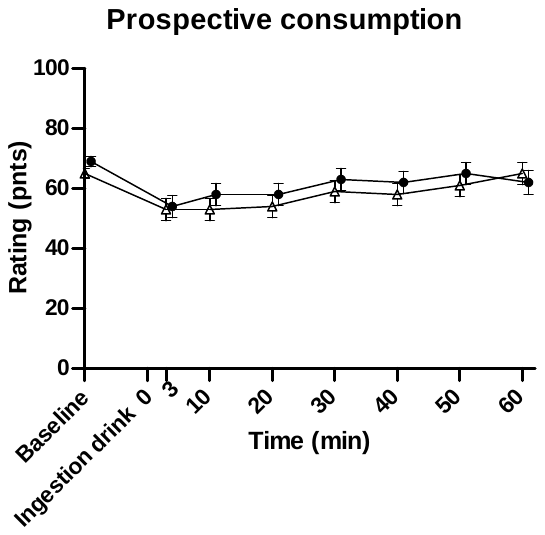 |
| 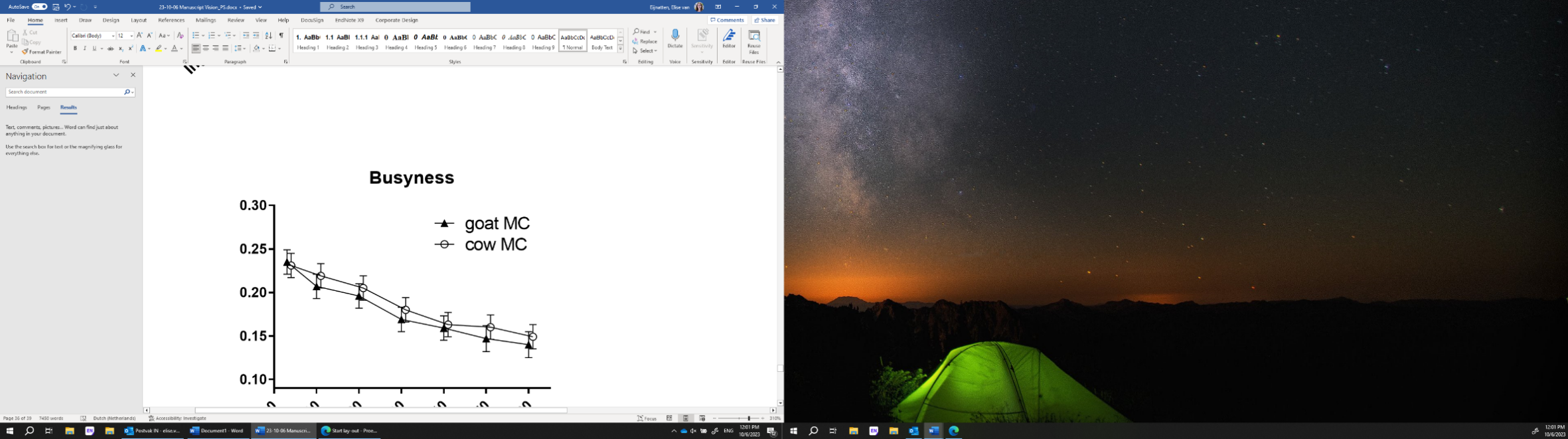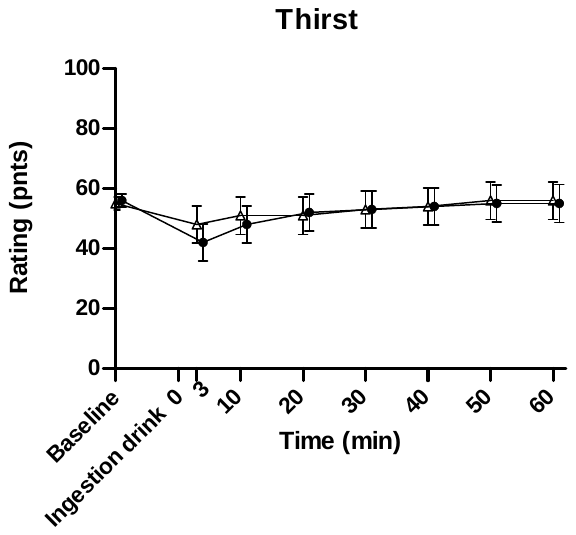 | |

***Supplemental figure 9.*** *Mean* ± *SEM appetite ratings, hunger, fullness, desire to eat over time after cow and goat milk-derived casein drink ingestion. *placed at the right of the graph it denotes a significant treatment effect (p<0.05). Above a data point it denotes a significant time point (post-hoc test p<0.05).*

| **Correlations** | | | | | | | | | | | |
| --- | --- | --- | --- | --- | --- | --- | --- | --- | --- | --- | --- |
|  | | GEt50 | Homogeneity_t30 | Contrast_t30 | Coarseness_t30 | Busyness_t30 | EAA | NEAA | BCAA | Glucose | Insulin |
| GEt50 | Pearson Correlation | 1 | ,337 | -,413^*^ | -,381^*^ | ,272 | -,159 | ,044 | -,108 | ,174 | ,329 |
|  | Sig. (2-tailed) |  | ,055 | ,017 | ,029 | ,126 | ,411 | ,822 | ,575 | ,316 | ,050 |
|  | N | 36 | 33 | 33 | 33 | 33 | 29 | 29 | 29 | 35 | 36 |
| Homogeneity_t30 | Pearson Correlation | ,337 | 1 | -,819^**^ | -,752^**^ | ,914^**^ | -,047 | -,052 | ,026 | -,135 | -,168 |
|  | Sig. (2-tailed) | ,055 |  | <,001 | <,001 | <,001 | ,814 | ,797 | ,896 | ,461 | ,350 |
|  | N | 33 | 33 | 33 | 33 | 33 | 27 | 27 | 27 | 32 | 33 |
| Contrast_t30 | Pearson Correlation | -,413^*^ | -,819^**^ | 1 | ,915^**^ | -,762^**^ | ,052 | -,025 | -,009 | ,231 | ,040 |
|  | Sig. (2-tailed) | ,017 | <,001 |  | <,001 | <,001 | ,796 | ,900 | ,964 | ,203 | ,824 |
|  | N | 33 | 33 | 33 | 33 | 33 | 27 | 27 | 27 | 32 | 33 |
| Coarseness_t30 | Pearson Correlation | -,381^*^ | -,752^**^ | ,915^**^ | 1 | -,624^**^ | -,090 | -,166 | -,144 | ,123 | -,004 |
|  | Sig. (2-tailed) | ,029 | <,001 | <,001 |  | <,001 | ,654 | ,408 | ,474 | ,501 | ,984 |
|  | N | 33 | 33 | 33 | 33 | 33 | 27 | 27 | 27 | 32 | 33 |
| Busyness_weighted_t30 | Pearson Correlation | ,272 | ,914^**^ | -,762^**^ | -,624^**^ | 1 | -,108 | -,091 | -,053 | -,221 | -,195 |
|  | Sig. (2-tailed) | ,126 | <,001 | <,001 | <,001 |  | ,591 | ,652 | ,792 | ,223 | ,276 |
|  | N | 33 | 33 | 33 | 33 | 33 | 27 | 27 | 27 | 32 | 33 |
| EAA_S | Pearson Correlation | -,159 | -,047 | ,052 | -,090 | -,108 | 1 | ,828^**^ | ,966^**^ | ,157 | ,302 |
|  | Sig. (2-tailed) | ,411 | ,814 | ,796 | ,654 | ,591 |  | <,001 | <,001 | ,424 | ,111 |
|  | N | 29 | 27 | 27 | 27 | 27 | 29 | 29 | 29 | 28 | 29 |
| NEAA_S | Pearson Correlation | ,044 | -,052 | -,025 | -,166 | -,091 | ,828^**^ | 1 | ,720^**^ | ,177 | ,491^**^ |
|  | Sig. (2-tailed) | ,822 | ,797 | ,900 | ,408 | ,652 | <,001 |  | <,001 | ,368 | ,007 |
|  | N | 29 | 27 | 27 | 27 | 27 | 29 | 29 | 29 | 28 | 29 |
| BCAA_S | Pearson Correlation | -,108 | ,026 | -,009 | -,144 | -,053 | ,966^**^ | ,720^**^ | 1 | ,128 | ,268 |
|  | Sig. (2-tailed) | ,575 | ,896 | ,964 | ,474 | ,792 | <,001 | <,001 |  | ,515 | ,160 |
|  | N | 29 | 27 | 27 | 27 | 27 | 29 | 29 | 29 | 28 | 29 |
| Glucose_AUC | Pearson Correlation | ,174 | -,135 | ,231 | ,123 | -,221 | ,157 | ,177 | ,128 | 1 | ,173 |
|  | Sig. (2-tailed) | ,316 | ,461 | ,203 | ,501 | ,223 | ,424 | ,368 | ,515 |  | ,319 |
|  | N | 35 | 32 | 32 | 32 | 32 | 28 | 28 | 28 | 35 | 35 |
| Insuline_AUC | Pearson Correlation | ,329 | -,168 | ,040 | -,004 | -,195 | ,302 | ,491^**^ | ,268 | ,173 | 1 |
|  | Sig. (2-tailed) | ,050 | ,350 | ,824 | ,984 | ,276 | ,111 | ,007 | ,160 | ,319 |  |
|  | N | 36 | 33 | 33 | 33 | 33 | 29 | 29 | 29 | 35 | 36 |
| *. Correlation is significant at the 0.05 level (2-tailed). | | | | | | | | | | | |
| **. Correlation is significant at the 0.01 level (2-tailed). | | | | | | | | | | | |

***Supplemental figure 10.*** *Correlations between image texture metrics at 30 min, 4-h AUC of blood parameters and gastric emptying half time*
